# Concurrent validity of an Estimator of Weekly Alcohol Consumption (EWAC) based on the Extended AUDIT

**DOI:** 10.1101/2020.12.11.20247106

**Authors:** PF Dutey-Magni, J Brown, J Holmes, JMA Sinclair

**Author notes:** **Corresponding author:** Peter F. Dutey-Magni, Institute of Health Informatics, University College London, 222 Euston Road, London, NW1 2DA, UK. **Competing interests** JB has received unrestricted research funding to study smoking cessation from companies who manufacture smoking cessation medications. PD, JH and JMAS declare no competing interests. **Protocol registration** Dutey-Magni, PF, Sinclair, JMA, Brown, J. 2018. ‘Concurrent Validity of an Estimator of Weekly Alcohol Consumption (EWAC) Based on the Extended AUDIT.’ OSF. November 12. doi:<u>10.17605/OSF.IO/7WE4M</u>.

## Abstract

**Background and Aims:** The 3-question Alcohol Use Disorders Identification Test (AUDIT-C) is frequently used in healthcare for screening and brief advice about levels of alcohol consumption. AUDIT-C scores (0–12) provide feedback as categories of risk rather than estimates of actual alcohol intake, an important metric for behaviour change. The study aimed to (a) develop a continuous metric from the Extended AUDIT-C expressed in United Kingdom (UK) units (8g pure ethanol), offering equivalent accuracy, and providing a direct estimator of weekly alcohol consumption (EWAC) and (b) evaluate the EWAC’s bias and error using the Graduated-Frequency (GF) questionnaire as a reference standard of alcohol consumption.

**Design:** Cross-sectional diagnostic study based on a nationally-representative survey.

**Settings:** Community-dwelling households in England.

**Participants:** 22,404 household residents aged ≥ 16 years reporting drinking alcohol at least occasionally.

**Measurements:** Computer-assisted personal interviews consisting of (a) AUDIT questionnaire with extended response items (the ‘Extended AUDIT’) and (b) GF. Primary outcomes were: mean deviation <1 UK unit (metric of bias); root mean squared deviation <2 UK units (metric of total error) between EWAC and GF. The secondary outcome was the receiver operating characteristic area under the curve for predicting alcohol consumption in excess of 14 and 35 UK units.

**Findings:** EWAC had a positive bias of 0.2 UK units [95% confidence interval: 0.08, 0.4] compared with GF. Deviations were skewed: while the mean error was ±11 UK units/week [9.5, 11.9], in half of participants the deviation between EWAC and GF was between 0 and ±2.1 UK units/week. EWAC predicted consumption in excess of 14 UK units/week with a significantly greater area under the curve (0.918 [0.914, 0.923]) than AUDIT-C (0.870 [0.864, 0.876]) or the full AUDIT (0.854 [0.847, 0.860]).

**Conclusions:** A new estimator of weekly alcohol consumption (EWAC), which uses answers to the Extended Alcohol Use Disorders Identification Test (Extended AUDIT-C), meets the targeted bias tolerance. It is superior in accuracy to AUDIT-C and the full AUDIT when predicting consumption thresholds, making it a reliable complement to the Extended AUDIT-C for health promotion interventions.

## Introduction

Alcohol consumption is responsible for 5% of disability-adjusted life years [1]. This burden extends far beyond the health burden of *alcohol use disorders*, as defined in the International Statistical Classification of Diseases (ICD-10 F10.1/F10.2 [2]) or the Diagnostic and Statistical Manual of Mental Disorders [3]. Clinical guidelines aiming to prevent [4], treat and reduce [5] harm from alcohol consumption recommend systematic screening for alcohol consumption using validated clinical tools. However, conceptual differences (exemplified by the diagnostic classifications above) remain in how best to diagnose, measure, and communicate harm [6].

A global standard has emerged in the 10-item Alcohol Use Disorders Identification Test (AUDIT) [7]. The shorter 3-item AUDIT-C focusses on consumption, and has equivalent predictive capability [8]. AUDIT-C is easy to use, making it an attractive choice for alcohol screening and brief interventions in healthcare [9] and other settings [10]. AUDIT-C exhibits two characteristics:

1. **Ceiling effect:** AUDIT-C’s maximum response options for alcohol consumption frequency and quantity are heavily right-censored (Table 1). This creates a ceiling effect making the AUDIT-C poorly responsive to change in individuals with a high baseline score (up to reductions of 30%; e.g. frequency of drinking down from 7 to 5 days or quantity down from 16 to 11 drinks per day).
2. **AUDIT score interpretation:** The ordinal scores produced by the AUDIT-C (range: 0–12) and the full AUDIT (range: 0–40) are multidimensional measures of alcohol risk. To date, most brief intervention models involve dichotomising AUDIT scores, on the basis of complex diagnostic accuracy studies [11], at cut-offs that vary internationally [12]. In practice, this may contribute to healthcare professionals lacking confidence in discussing alcohol risks and consumption [13–16], and needing to be trained to deliver feedback [13,14]. Evidence also suggests that patients’ understanding of alcohol risks overlaps loss of control more than alcohol consumption [17,18]. In response, some academic models of alcohol care advocate the framing of brief interventions around the continuum of alcohol use [19] rather than thresholds, since these can trigger stigma related to loss of control [20].

**Table 1.** Comparison of AUDIT-C and Extended AUDIT-C

| Response items |  |  |  |  |  |  |  |
| --- | --- | --- | --- | --- | --- | --- | --- |
| AUDIT-1: 'How often do you have a drink containing alcohol?' |  |  |  |  |  |  |  |
| AUDIT-C | Never | Monthly or less | 2 - 4 times per month | 2 - 3 times per week | 4+ times per week | – | – |
| Extended AUDIT-C | Never | Monthly or less | 2 - 4 times per month | 2 - 3 times per week | 4-5 times per week | 6+ times per week | – |
| [score] | [0] | [1] | [2] | [3] | [4] | [4] | – |
| AUDIT-2: 'How many units of alcohol do you drink on a typical day when you are drinking?' |  |  |  |  |  |  |  |
| AUDIT-C | 0 - 2 | 3 - 4 | 5 - 6 | 7 - 9 | 10+ | – | – |
| Extended AUDIT-C | 0 - 2 | 3 - 4 | 5 - 6 | 7 - 9 | 10-12 | 13-15 | 16+ |
| [score] | [0] | [1] | [2] | [3] | [4] | [4] | [4] |
| AUDIT-3: 'How often have you had 6 or more units on a single occasion in the last year?' |  |  |  |  |  |  |  |
| AUDIT-C | Never | Monthly or less | Monthly | Weekly | Daily or almost daily | – | – |
| Extended AUDIT-C | Never | Monthly or less | Monthly | Weekly | Daily or almost daily | – | – |
| [score] | [0] | [1] | [2] | [3] | [4] | – | – |

The ‘Extended AUDIT-C’ addresses the first characteristic thanks to a greater range of response options on quantity and frequency (Table 1). It has been used in the United Kingdom (UK) as part of two trials [21,22] and one continuous household survey [23] to measure characteristics of consumption that could not have been measured with the right-censored AUDIT-C.

The present study proposes to address the second of these characteristics. It aims to develop and validate an Estimator of Weekly Alcohol Consumption (EWAC) computed from the Extended AUDIT-C. While retaining the Extended AUDIT-C questionnaire’s alcohol use disorders diagnostic capabilities, the EWAC is intended to facilitate the delivery of screening and brief interventions by converting Extended AUDIT-C responses into a continuous and direct measure of alcohol consumption that does not require additional screening questions. Measuring alcohol consumption is a crucial part of behaviour change techniques (self-monitoring, feedback on behaviour, social comparison) commonly employed in self-[10] and clinician-administered [9,24] interventions, and is encouraged as a metric of the continuum of alcohol use [19].

## Methods

### Participants

Data originate from baseline measures in waves 110–133 (November 2015–October 2017) of the Alcohol Toolkit Study, a repeated cross-sectional survey of residents of private English households aged ≥ 16 years. Each month, census output areas averaging 300 households were selected by stratified random sampling. Interviewers travelled to their designated area and approach households quota sampling [23]. Respondents participated in a computer-assisted personal interview.

### Measures

Index measurements underpinning the EWAC were the three questions making up the Extended AUDIT-C (supplementary information S2), in which participants described their drinking *during the last 6 months*.

The reference standard used is the Alcohol Toolkit Study GF schedule (supplementary information S3), in which participants described how many times they consumed given quantities of alcohol *during the last 4 weeks* [25]. The GF schedule’s main advantage lies in measuring occasional heavy consumption, which can constitute an important proportion of total consumption.

Other reference estimates were used, this time for aggregate comparisons. 2014 per-capita alcohol retail sales [26] captured all alcohol produced/processed in or imported to England for sale or consumption. We also used data from 6,606 household residents aged ≥ 18 years participating in the 2011 Health Survey for England [27]. Year 2011 was chosen in deviation from the registered protocol [28]: on that particularly year, the recurring computer-assisted interviewer-led beverage-specific quantity-frequency questionnaire was accompanied by a prospective 7-day diary [29]. The diary reference standard was deemed more informative to an international audience, and offered a direct point of comparison with past research [30–32].

### Estimating alcohol consumption (EWAC)

To estimate alcohol consumption from Extended AUDIT-C responses, we employ methods developed for quantity-frequency-variability instruments [33]. For every individual *i*, the EWAC is computed as the product of *F*_*i*_ and *Q*_*i*_ (AUDIT questions 1 and 2 respectively) adjusted with the frequency of intense drinking *V*_*i*_ (AUDIT-3):

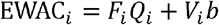

where *b* denotes the average units of alcohol consumed in an intense drinking day.

Coefficients *F, Q, V* and *b* are unknown. In this study, two sets of candidate coefficients are considered:

- AUDIT response item interval midpoint (e.g. 2.5 for ‘2 to 3 times per week’)
- coefficients estimated empirically from a sample of individuals with measurements of Extended AUDIT-C and GF, using a hierarchical Bayesian response model with the estimating equation GF_*i*_ = *F*_*i*_ *Q*_*i*_ + *vib*+ *e*_*i*_, where *e* denotes independently normally distributed errors. We set parabola-shaped informative priors on coefficients *F, Q, V*. Details on model fitting, convergence evaluation and prior tuning are reported in supplementary information S1.

### Analyses

The protocol was pre-registered [28]. Results are reported in UK alcohol units (8g or 10mL of pure alcohol). Analyses were conducted in R [34–36] and all computer scripts are available online [37].

Participants were included in the analysis if they completed both the Extended AUDIT and the GF questionnaires. Out of 40,832 participants, 14,408 (35%) reported ‘never’ consuming alcohol in AUDIT question 1 and were not asked any further AUDIT or GF questions. A further 175 (0.4%) did not have valid AUDIT-C answers. Finally, 3,876 participants (9%) who did not have a valid GF alcohol consumption record were excluded. These GF data were assumed to be missing at random conditionally on the Extended AUDIT-C responses after a sensitivity analysis (supplementary information S1).

Valid observations (*N*=22,373) were separated into two datasets:

- The training dataset (*N*=6,642) consisted of a 30 percent subset of participants drawn using stratified random sampling, ensuing a balanced representation by sex, age, ethnic group and AUDIT-C risk level. It was used to estimate coefficients underpinning the EWAC (supplementary information S1).
- The validation dataset consisted of the remaining participants (*N*=15,731) and was used to evaluate the EWAC’s bias and precision. In subgroup validation analyses utilising additional variables (eg education, smoking status), a further 358/15,731 observations (2.3%) assumed to be missing at random were excluded.

#### Overall bias and error

The agreement between the EWAC and the GF was quantified in the validation dataset:

- bias was estimated by the **mean deviation** to the reference standard 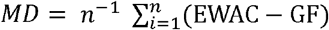. We tested the hypothesis that the MD does not exceed 1 UK unit using a two-sided t-test.
- precision was estimated by the **root mean squared deviation** 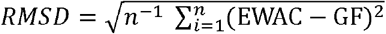, a measure of total error capturing both bias and random deviation from the reference standard. For example, an RMSD of 2 signifies that the EWAC is on average with ± 2 UK units of the reference standard. We tested the hypothesis that the RMSD does not exceed 2 UK units using a one-sided χ^2^ test.

Two sets of candidate coefficients were considered (see ‘Design’ section). We only report findings for the candidate set producing the lowest bias and error.

#### Subgroup bias and error

Multivariate regression models tested whether the EWAC’s bias and precision varied across population subgroups:

- the simple deviation (EWAC - GF) was regressed in a linear model to test subgroup differences in MD
- the squared deviation (EWAC - GF)^2^ was regressed in a log-transformed linear model to test subgroup differences in the geometric mean squared deviation. Model coefficients were then back-transformed (square root of the exponential) into relative RMSD estimates; these are interpreted as the ratio of the subgroup RMSD to the reference RMSD, a ratio >1 indicating worse precision than in the reference category.

Both models (supplementary table S5.1) included the following predictors: sex by age group; ethnic group; highest educational qualification; religion; smoking status. Additional models (supplementary table S5.2) were fitted solely in respondents with an AUDIT-C score ≥ 5 or an AUDIT score ≥ 8, for whom additional characteristics were recorded during interview: favourite drink (beer; wine; spirits alone; mixed spirits; cider; other); and whether the respondent had attempted to restrict alcohol intake in the last 12 months.

#### Receiver Operating Characteristics

We tested the EWAC’s superiority to the traditional AUDIT and AUDIT-C scores in predicting consumption exceeding 14 or 35 UK units/week. These correspond to UK thresholds for characterising alcohol use as ‘increasing risk’ (predicted by an AUDIT-C score of 5–7), and ‘higher risk’ (AUDIT-C score ≥ 8) which is above 35 units for women and 50 units for men [38]. We tested the hypothesis that the EWAC has an identical receiver operating characteristic full area under the curve (AUC) to the AUDIT-C and the full AUDIT scores using nonparametric paired AUC tests [39]. AUDIT-C and AUDIT scores were calculated from the Extended AUDIT by capping the contribution of each question to 4.

#### Aggregate concurrent validity

We compared the empirical cumulative distributions of (1) the EWAC computed in the Alcohol Toolkit Study; (2) the GF estimator in the Alcohol Toolkit Study; (3) the beverage-specific estimator in the 2011 Health Survey for England; (4) the prospective diary estimator in the 2011 Health Survey for England in adults aged ≥ 18 years. A χ^2^ test of homogeneity of distributions (1) and (3) was performed on contingency tables of 13 drinking consumption intervals in UK units/week (]0,5]; ]5,10]; …; ]30,35]; ]35,45]; ]45,55]; ]55,65]; ]65,75]; ]75,100]; ]100,200]). We report the proportions of on-trade and off-trade alcohol sales [26] accounted for by each method. Poststratification survey weights adjusted for nonresponse bias in sources (1-3), and self-selection into prospective diary data collection in source (4).

## Results

### Bias and precision

EWAC coefficients estimated empirically (supplementary information S1, S4) had smaller bias and error and were used for the remainder of the analysis. With those, the EWAC’s Pearson’s correlation with GF was estimated at *r* = 0.72 [0.71, 0.72] (Kendall’s rank correlation *τ* = 0.63).

The mean deviation (MD) was 0.2 alcohol units/week [95% CI: 0.08, 0.4]. This bias is smaller than the preregistered ± 1-unit bias tolerance (p = 1.000).

The root mean squared deviation (RMSD), at 10.7 units/week [95% CI: 9.5, 11.9], was significantly greater than the pre-registered 2-unit total error tolerance (*p* < 0.001), suggesting that the EWAC falls on average 11 units away from the GF reference standard.

However, there was substantial variation in RMSD; in 50% of participants, the EWAC fell within ± 2.1 UK units of the GF weekly consumption estimate. RMSD was proportional to alcohol consumption, amounting to about 50% of the EWAC value (Table 2). Thus, an interval defined as the EWAC ± 50% (e.g. ‘2–6 units/week’ for an EWAC of 4; ‘10–30 units/week’ for an EWAC of 20) contained the reference standard for over half (58%) of individuals.

**Table 2.** Root Mean Squared Deviation (RMSD) between EWAC and GF schedule by alcohol consumption bracket (n = 15,731)

| EWAC value<br>(UK units/week) | n | RMSD<br>[95% confidence<br>interval] | Participants with<br>GF contained in<br>[EWAC×0.5; EWAC×1.5]<br>interval (%) |
| --- | --- | --- | --- |
| [0,5) | 6,927 | 3.1 [2.7–3.5] | 3,375 (48.7) |
| [5,10) | 3,589 | 10.0 [3.8–13.5] | 2,127 (59.3) |
| [10,20) | 3,363 | 12.4 [9.3–14.9] | 2,330 (69.3) |
| [20,30) | 1,010 | 15.5 [12.6–17.9] | 736 (72.9) |
| [30,45) | 495 | 19.4 [17.0–21.4] | 342 (69.1) |
| [45,60) | 142 | 27.4 [22.1–31.8] | 101 (71.1) |
| [60,75) | 113 | 25.7 [18.9–31.0] | 95 (84.1) |
| [75,100) | 66 | 40.2 [25.1–51.0] | 48 (72.7) |
| [100,150) | 26 | 77.0 [59.2–91.4] | 13 (50.0) |
| **All values** | 15,731 | 10.7 [9.4–11.9] | 9,167 (58.3) |

Plots of EWAC against GF (Figure 1) indicate a slight positive bias for consumptions up to 10-14 units/week, and a slight negative bias beyond. The EWAC only starts losing granularity above 70 units/week (99th percentile of its distribution), where it provides just 6 possible values (82; 83; 92; 93; 100; 125 units/week; see Figure 1(b)).

**Figure 1.**
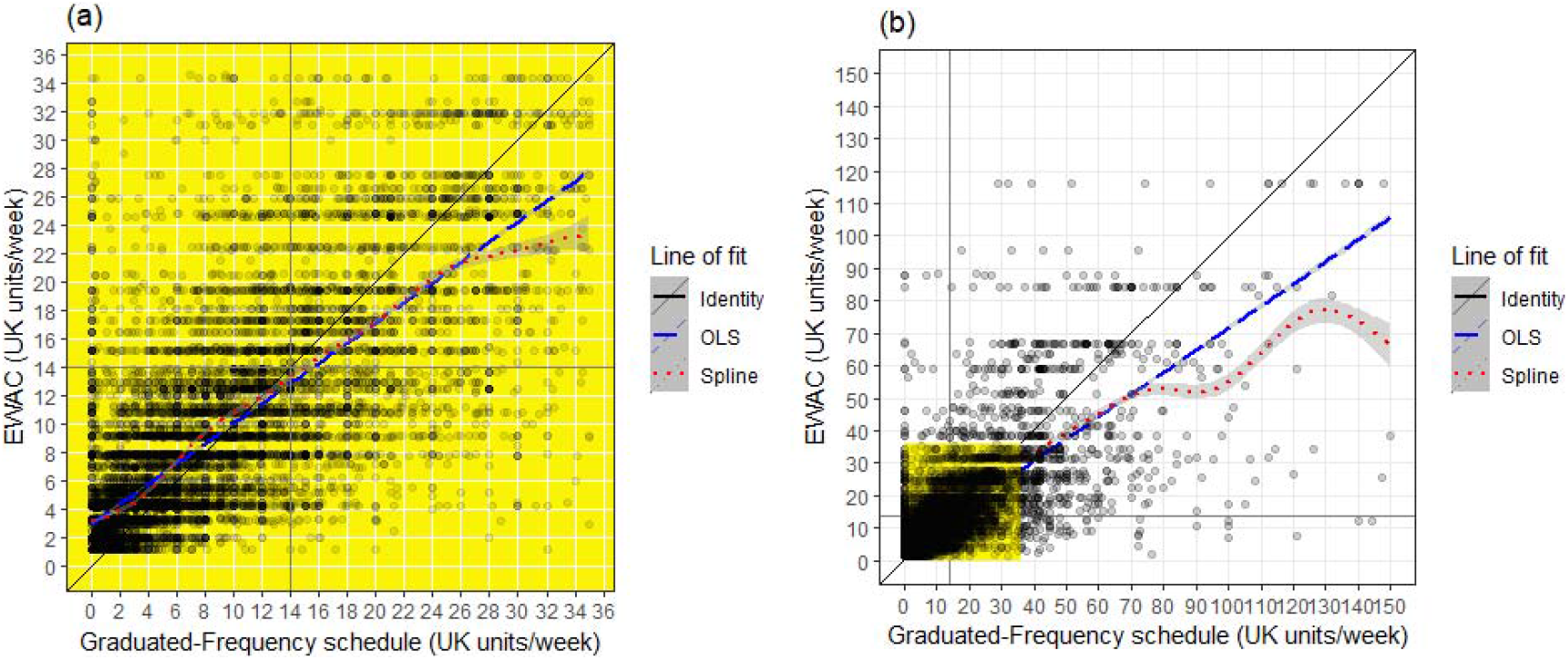
Plots of EWAC against GF in (a) low/increasing risk respondents (n=15,008) and (b) all respondents (n=15,731)

Extensive subgroup analyses are reported in supplementary material 5. A very modest proportion of variation in bias and precision (<5%) can be attributed to socio-demographic variables under examination. This indicates a relative homogeneity in precision in bias, to one exception. The EWAC appears to overestimate consumption by 1 to 2 UK units in groups with the lowest average consumption: women, and Non-British White, Black, and Other ethnic groups.

### Receiver Operating Characteristics

We examined the EWAC’s ability to predict consumption exceeding 14 or 35 UK units/week. The full areas under the receiver operating characteristic curves (AUC, supplementary Figure S6) are presented along sensitivity and specificity at the best thresholds in Tables 3– 4.

**Table 3.** Receiver operating characteristics of AUDIT-C score and EWAC for consumption >= 14 UK units or 112g/week (n = 15,731)

| Index test | Full Area Under the Curve | 95% CI | Best threshold | Sensitivity | Specificity |
| --- | --- | --- | --- | --- | --- |
| AUDIT-C score | 0.870 | [0.864, 0.876] | 5.5 | 0.753 | 0.811 |
| Full AUDIT score | 0.854 | [0.847, 0.860] | 5.5 | 0.792 | 0.751 |
| EWAC | 0.918 | [0.914, 0.923] | 9.8 | 0.873 | 0.813 |

*Note:* The best threshold refers the cut-off value that maximises the sum of sensitivity and specificity.

**Table 4.**
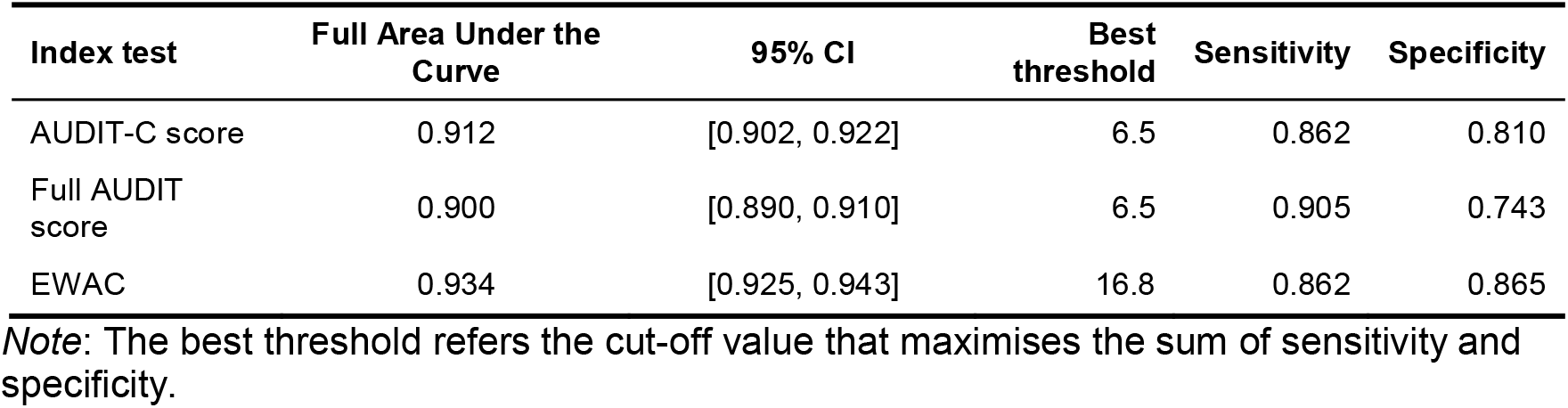
Receiver operating characteristics of AUDIT-C score and EWAC for consumption >= 35 UK units or 280g/week (n = 15,731)

| Index test | Full Area Under the Curve | 95% CI | Best threshold | Sensitivity | Specificity |
| --- | --- | --- | --- | --- | --- |
| AUDIT-C score | 0.912 | [0.902, 0.922] | 6.5 | 0.862 | 0.810 |
| Full AUDIT score | 0.900 | [0.890, 0.910] | 6.5 | 0.905 | 0.743 |
| EWAC | 0.934 | [0.925, 0.943] | 16.8 | 0.862 | 0.865 |

*Note:* The best threshold refers the cut-off value that maximises the sum of sensitivity and specificity.

#### 14 UK units/week (increasing-risk)

EWAC increases the AUC by 5 percentage points compared to the AUDIT-C score (*p* < 0.001); and 7 percentage points compared to the full AUDIT score (*p* < 0.001). The cut-off maximising the sum of specificity and sensitivity on the EWAC is 10 units/week. The sensitivity at this threshold is identical to AUDIT-C, but specificity gains 13 percentage points. Using the nominal cut-off of 14 units/week on the EWAC raises specificity to 0.928, at the cost of a reduction in sensitivity to 0.687 (Table 3).

#### 35 units/week (higher-risk)

EWAC provides small increases in AUC compared with the AUDIT-C score (*p* < 0.001) and the full AUDIT score (*p* < 0.001). The best cut-off for detecting consumption in excess of 35 units/week using the EWAC was 17 units/week (Table 4).

### Empirical distribution

Table 5 estimates adult residents’ total alcohol consumption in England using four different sources, and compares them with alcohol retail sales. The Health Survey for England exhibits the highest estimates and coverage of alcohol sales. EWAC amounts to 71% of the total consumption estimated by the Health Survey for England’s prospective diary, and 48% of retail sales.

**Table 5.** Summary statistics on alcohol consumption in England in residents aged 18 years and over (excluding abstainers)

| Study | Mean (UK units/week) | Median (UK units/week) | Variance | N | % of alcohol sold |
| --- | --- | --- | --- | --- | --- |
| HSE beverage-specific QF | 14.0 | 7.3 | 474.6 | 6,545 | 72.6 |
| HSE prospective diary | 13.0 | 8.0 | 264.7 | 4,640 | 67.6 |
| ATS GF | 8.5 | 5.2 | 234.6 | 15,556 | 43.9 |
| ATS EWAC | 9.3 | 5.2 | 145.9 | 18,140 | 48.2 |
| **Retail sales** | 19.3 | – | – | – | 100.0 |

Figure 2 suggests that the EWAC, like the Alcohol Toolkit Study GF, estimates a greater prevalence of lower-risk (≤ 14 units/week) and increasing-risk alcohol use than Health Survey for England. It shows a clear departure between the EWAC and the Health Survey for England’s beverage specific questionnaire, as evidence by the homogeneity test 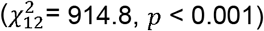.

**Figure 2.**
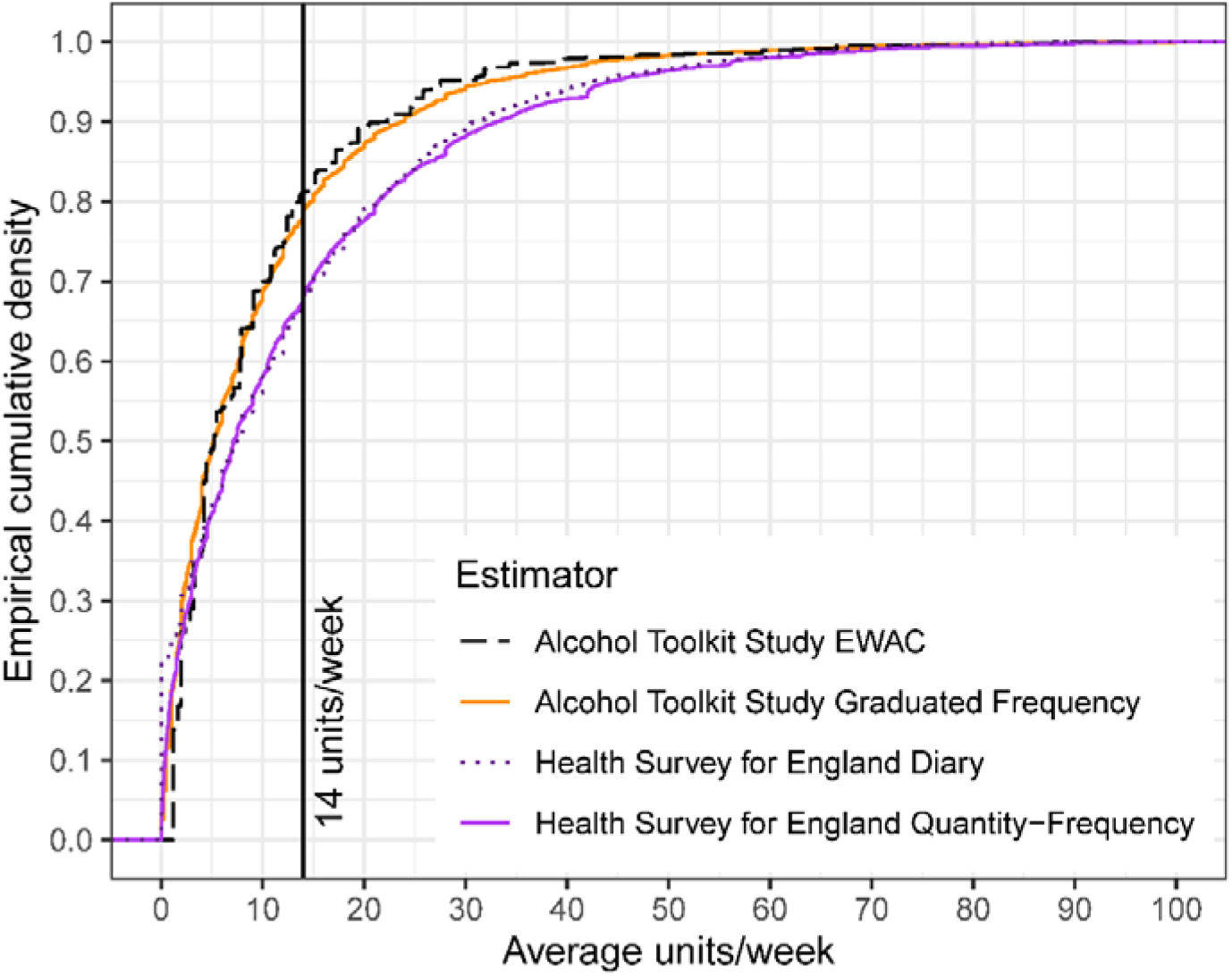
Empirical cumulative distribution function of weekly alcohol consumption in England according to four alcohol schedules in residents aged 18 years and over

## Discussion

### Main findings

We developed a continuous Estimator of Weekly Alcohol Consumption (EWAC) using a 6-month Extended AUDIT-C. When compared with a 4-week Graduated Frequency (GF) reference standard, we found EWAC overestimates alcohol consumption by 0.2 UK units [0.08, 0.4], well under the pre-registered ±1 UK unit bias tolerance. We also attempted to measure how precise the EWAC is: in 50% of participants, the EWAC falls up to 2 UK units away from the GF measure, and an interval built as EWAC ±50% contains the GF measures in 58% of participants.

EWAC is superior to both the AUDIT-C and the full AUDIT scores in predicting GF exceeding 14 units/week (AUC = 0.92) and 35 units/week (AUC = 0.93). This places the EWAC among the best-performing diagnostic tools examined in the most recent systematic review [11]. At the 14-unit threshold, an EWAC >= 10 cut-off has a sensitivity of 0.87, compared to a 0.75 for an AUDIT-C >= 6 cut-off, without losing specificity.

### Potential applications

Being equivalent to the AUDIT-C in speed and international standardisation, the EWAC may be suitable for use in any clinical setting to support brief interventions and to feed back a reliable interval estimate of alcohol consumption (eg: ‘6–18 units/week’ or ‘50–140g/week’). The EWAC is available as a web app at https://ewac.netlify.app along with resources to facilitate implementation (R software package, spreadsheets).

Assessment of alcohol consumption is not well embedded in clinical practice [40]. The EWAC calculator fills a gap in resources by transforming the answers from the Extended AUDIT-C into a direct estimate of an individual’s weekly alcohol consumption. This is a more directly accessible metric which should facilitate behaviour change by empowering people to monitor and control their alcohol consumption with–or without–the involvement of healthcare professionals, and should be assessed in future evaluations.

Nutt, Rehm *et al*. [19,41] argued that alcohol-related harm is best prevented if individuals know their consumption level, and health professionals in all settings can engage patients effectively to manage risks with evidence-based interventions, in a similar way to other risk factors for disease, for example blood pressure or cholesterol. Yet, knowledge of beverages’ alcohol content is generally poor [42], and a survey evaluating the 2016 change in UK alcohol guidelines found that just 8% of the UK drinkers knew the new recommended limits [43]. The EWAC can support interventions focused on recognising the alcohol content/volume of drinks, and recommended low-risk limits.

In addition, the EWAC’s dimensional rather than categorical format can be useful to position recipients of brief interventions on the continuum of alcohol use [20,44], which may reduce the stigma of loss of control associated with screening-based interventions [17,45]. It can act as a complement, rather than a substitute to the multidimensional quality of AUDIT-C or the full AUDIT.

The EWAC is particularly suitable for digital interventions and healthcare records given that it enables its complex algorithm to be embedded in a way not possible with paper records. EWAC is already compatible with medical records information models developed in the Systematised Nomenclature of Medicine Clinical Terms (SNOMED CT, Alcohol intake (observable entity) [46]) and by the English Royal College of Physicians [47]. Such information can have secondary uses as a variable in other disease risk scores, or to prospectively recording of long-term alcohol exposure, an important risk factor for a range of medical conditions.

### Strengths and limitations

This paper is the first to (a) develop an EWAC using a well-accepted and validated multidimensional alcohol screening tool such as the AUDIT; and (b) quantify its bias and precision with respect to a continuous measure of alcohol consumption. One study [48] previously reported mean consumption by AUDIT-C score, but without quantifying bias or precision of such a measure. Others have evaluated the AUDIT-C’s accuracy in estimating alcohol consumption, but exclusively in relation to predicting consumption in excess of predefined thresholds [11].

Our study provides strong confidence in the internal and external validity of findings in England on account of the large sample size and extensive range of subgroup analyses reported. Bias was mostly consistent across subgroups examined (age/sex, education, smoking status, religion), with one exception. EWAC overestimated alcohol consumption by 2-3 UK units/week in Black/Other ethnic groups. Variation in the sensitivity of AUDIT-C across ethnic groups has previously been noted in the US [49].

Repurposing a well-known tool such as the AUDIT-C has several advantages. It is already translated in many languages and adapted to the varying standard drink sizes adopted internationally [7]. The Extended AUDIT scores can be converted into traditional AUDIT scores by capping items to 4, thereby offering a point of comparison with existing evidence. The AUDIT’s properties are also well understood in diverse contexts and modes of administration, based on the last 30 years of international research. For instance, a previous study which found the AUDIT-C to be responsive to changes of 70g/week [50] can suggest that the EWAC’s own responsiveness to change should be equivalent, if not greater than the AUDIT-C’s, given the Extended AUDIT-C’s additional response items.

We note two main study limitations. First, a longstanding obstacle in alcohol research and treatment lies in the absence of undisputed ‘gold standard’ or biomarker for objectively determining alcohol consumption. Instead, a number of instruments measure self-reported consumption with varying validity and reliability over different durations. Comprehensive reviews [30–32,51–53] indicate that yesterday recall and prospective diaries tend to record higher (and more accurate) alcohol consumption by minimising recall bias, followed by GF measures.

Therefore, the GF reference standard, as all self-reported measures, is imperfect. While this has no effect on our measure of bias (MD), this may introduce bias into our measure of precision (RMSD): by definition, the reference standard’s own independent error will inflate the RMSD. In other words, it is likely that a proportion of the RMSD is attributable to error in the GF measures rather than the EWAC.

Despite this, previous research suggests the EWAC’s agreement with GF (Pearson’s correlation coefficient *r* = 0.71 and Kendall’s rank correlation *r* = 0.63 in the present study) is comparable to the agreement between GF and prospective diaries measured from past studies(*r* ∼ 0.86–0.89 [54,55]; *τ* = 0.41 [51]).

Second, the EWAC’s design does not escape all limitations of methods of screening or categorising alcohol use disorders. The conceptualisation of alcohol use disorders is related to, but does not exclusively depend upon the amount of alcohol consumed. Since Jellinek’s description of ‘the disease concept of alcoholism’ [56] there have been numerous attempts to categorise the range of phenotypes characterising alcohol use disorders in the absence of any biomarker to ‘verify’ the presence of a particular pathology. The EWAC, by limiting itself to an estimation of alcohol consumption is transparent across a wide range of alcohol use disorders but does not measure the other factors underpinning this complex and heterogeneous condition [6,57].

In conclusion, the EWAC has the potential to support interventions focusing on recognising the alcohol content and volume of drinks. The EWAC’s dimensional rather than categorical format may facilitate this while avoiding the stigma sometimes associated with clinical categorisations of alcohol use disorders.

## Supporting information

S1: Model specifications

S2: ATS AUDIT questionnaire

S3: ATS Graduated Frequency questionnaire

S4: EWAC coefficients

S5: Subgroup analyses

S6: ROC curves

S7: Demographics

S8: STARD checklist

## Data Availability

ATS data can be requested from the Alcohol Toolkit Study team.

https://doi.org/10.5281/zenodo.4315023

## Declarations

## Acknowledgements

This research was supported by the Medical Research Council [grant reference MR/P016960/1]. The Alcohol Toolkit Study data collection was funded primarily by the National Institute for Health Research (NIHR) School for Public Health Research [grant reference SPHR□SWP□ALC□WP5] and Public Health Research Programme [grant reference 15/63/01]. The EWAC online calculator development was funded by the Wessex Academic Health Science Network. The views expressed are those of the author(s) and not necessarily those of the NHS, the NIHR or the Department of Health.

## Ethics

This study was approved by the University of Southampton’s Faculty of Medicine Ethics Committee (ERGO 44682).

## Consent for publication

Not applicable.

