## Supplementary material for "Concurrent validity of an Estimator of Weekly Alcohol Consumption (EWAC) based on the Extended AUDIT": S1: Model specifications

**Supplementary information S1:
Model-based estimation of EWAC coefficients**

### 1 Data source and missingness

The Alcohol Toolkit Study (ATS) dataset provides both Graduated Frequency (GF) and Extended AUDIT-C measurements for a sample of household residents in England between waves 110 and 133, that is, November 2015 and October 2017. We restrict the analysis to adults aged 16 years and over. The GF schedule was not administered to participants responding 'Never' in AUDIT item 1. Consequently, this analysis is conducted only on drinkers (n = 22,373).

A model was fitted on the dataset with respondents completing the full Extended AUDIT-C (*n*= 26,249; excluding those answering AUDIT-1 with ‘Never’), entering AUDIT-C answers, the EWAC, and socio-demographic variables commonly associated with either nonresponse or alcohol consumption. Table 1 reports the most parsimonious model, after testing for some interactions and removing variables inflating the Akaike Information Criterion. We find evidence that the Graduated Frequency data is missing not at random.

To examine the influence of GF data missingness on our validation data, we performed a sensitivity analysis using multiple imputation with chained equations (MICE [1]) on the primary outcomes of the study: mean deviation and root mean square deviation in the validation sample. Using 20 imputations and default imputation methods (predictive mean matching for GF, polytomous regression imputation for the remaining categorical variables), we found:

- MD = 0.19 UK units/week with imputation, compared to 0.24 without imputation.
- RMSD = 10.38 UK units/week with imputation, compared to 10.73 without imputation.

These results being similar, we made the assumption that GF was Missing At Random Conditionally on Extended AUDIT-C answers.

Table 1 Logistic model of Graduated-Frequency missingness

|  |  | *Odds Ratios* | *Std. Error* | *p-value* |
| --- | --- | --- | --- | --- |
| (Intercept) |  | 0.721 | 0.248 | 0.341 |
| EWAC |  | 1.025 | 0.006 | **<0.001** |
| AUDIT-1 | Monthly or less | *Reference* |  |  |
|  | 2 to 4 times a month | 0.171 | 0.009 | **<0.001** |
|  | 2 to 3 times a week | 0.120 | 0.008 | **<0.001** |
|  | 4 to 5 times a week | 0.078 | 0.010 | **<0.001** |
|  | 6 or more times a week | 0.063 | 0.010 | **<0.001** |
| AUDIT-2 | 1 to 2 | *Reference* |  |  |
|  | 3 to 4 | 0.811 | 0.044 | **<0.001** |
|  | 5 to 6 | 0.879 | 0.073 | 0.118 |
|  | 7 to 9 | 0.799 | 0.085 | **0.035** |
|  | 10 to 12 | 0.817 | 0.108 | 0.127 |
|  | 13 to 15 | 1.273 | 0.255 | 0.229 |
|  | 16 or more | 1.023 | 0.168 | 0.891 |
| AUDIT-3 | Never | *Reference* |  |  |
|  | Less than monthly | 0.675 | 0.039 | **<0.001** |
|  | Monthly | 0.407 | 0.039 | **<0.001** |
|  | Weekly | 0.373 | 0.049 | **<0.001** |
|  | Daily or almost daily | 0.226 | 0.089 | **<0.001** |
| Marital status | Married | *Reference* |  |  |
|  | Separated/widowed | 1.108 | 0.066 | 0.085 |
|  | Single | 1.211 | 0.067 | **0.001** |
| Smoking | Never smoked | *Reference* |  |  |
| status | Stopped>1y ago | 1.144 | 0.061 | **0.012** |
|  | Stopped in past year | 0.906 | 0.153 | 0.560 |
|  | Smoker | 1.242 | 0.068 | **<0.001** |
| Sex | Women | *Reference* |  |  |
|  | Men | 0.877 | 0.037 | **0.002** |
| Age | 16-17 years | *Reference* |  |  |
|  | 18-24 years | 1.232 | 0.411 | 0.532 |
|  | 25-34 years | 1.090 | 0.366 | 0.798 |
|  | 35-44 years | 1.191 | 0.401 | 0.603 |
|  | 45-54 years | 0.997 | 0.336 | 0.994 |
|  | 55-64 years | 1.101 | 0.370 | 0.775 |
|  | 65-74 years | 1.176 | 0.397 | 0.630 |
|  | 75-84 years | 1.200 | 0.410 | 0.593 |
|  | 85+ years | 1.298 | 0.465 | 0.467 |
| Tenure | Owned | *Reference* |  |  |
|  | Rented | 1.464 | 0.069 | **<0.001** |
| Ethnic group | White British | *Reference* |  |  |
|  | White Other | 1.077 | 0.086 | 0.353 |
|  | Mixed | 0.818 | 0.144 | 0.254 |
|  | Asian | 1.136 | 0.150 | 0.335 |
|  | Black | 1.083 | 0.133 | 0.514 |
|  | Other | 0.650 | 0.155 | 0.070 |
| Highest | No qualification | *Reference* |  |  |
| qualification | NVQ <= 3 | 0.716 | 0.045 | **<0.001** |
|  | NVQ4+ (degree) | 0.563 | 0.040 | **<0.001** |
|  | Other | 0.890 | 0.079 | 0.190 |
| Religion | No religion | *Reference* |  |  |
|  | Christian | 0.892 | 0.040 | **0.011** |
|  | Muslim | 1.900 | 0.463 | **0.009** |
|  | Any other religion | 1.135 | 0.115 | 0.213 |
| Survey weights | | 1.041 | 0.058 | 0.478 |
| Observations: 25,268 | | AIC: 16,880 | | |

### 2 Evaluation of midpoint-based coefficients

Before taking a model-based approach, we tested coefficients based on AUDIT response item interval midpoints (e.g. 2.5 for '2 to 3 times per week') in the validation dataset (*n* = 15,731). Using those, the mean deviation (MD, measure of bias) was 0.7 UK units/week not significantly greater than 1 (*p* = 0.999). The root mean squared deviation (RMSD, measure of precision) was 12.1 UK units/week and significantly greater than 2 (p < 0.001).

### 3 Estimation of model-based coefficients

A 30 percent training dataset (*n* = 6,642) was selected using stratified random sampling, to ensure this dataset was balanced representation across the following strata: sex × ethnic group (White British; White Other; Non-White) × age (4 groups) × AUDIT-C risk level (Low, Increasing; High).

We employed a Bayesian response model with the following estimating equation

$$\mu_{i}=F_{i}Q_{i}+V_{i}b$$

$$\mathrm{GF}_{i} \sim\mathrm{Normal}\left( \mu_{i}, \sigma\right)$$

$$\sigma\sim\mathrm{Exponential}(0.1)$$

$GF$ is modelled in UK units/week. $F$ and $V$ were defined as weekly frequencies, while $Q$ and $b$ were defined in UK units (8g).

The model was estimated with 3,000 burn-in iterations, followed by 5,000 iterations that were kept as posterior estimates. Three independent Markov chains were used.

Prior tuning is an iterative process informed by patterns in convergence. The complete history of priors used is available from the version history of the ‘03_estimate_weights/stan_model.stan’ in the [GitHub repository](https://github.com/peterdutey/ewac-validation). Comments can be found in git commit messages as well as ‘findings.txt’ files under respective directories stored under ‘03_estimate_weights’.

We set starting informative priors on coefficients $F,Q,V$ in the form of uniform priors bounded by the response item interval and centred on its midpoint. For instance, the prior for AUDIT-Q2 *‘1 to 2’* alcohol units, was set to $Q_{2} \sim\mathrm{Uniform}(1, 2.5)$, and AUDIT-Q2 *‘3 to 4’* units to $Q_{2} \sim\mathrm{Uniform}(2.5, 4.5)$. Convergence was examine to verify that such intervals were realistic given the data, by looking at chains that all converged to the extreme values of the interval. This was the case for a number of parameters, including AUDIT-Q2 ‘13 to 15’ (Figure 1), which consistently converged to the bottom of the interval and was gradually brought to a lower prior bounded to [9.5; 15.5].

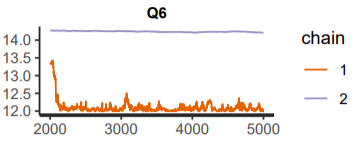

Figure 1 Initial model traces for AUDIT-Q2 ‘13 to 15’ units showing a pattern of convergence to the lower bound of the interval prior

Such priors were gradually tuned one after the other by updating interval priors. As convergence improved and chains converged to form healthy bell-shape posteriors, uniform priors were altered to parabola-shaped priors based on the $\mathrm{Beta}(2,2)$ hyperprior distribution. This was to make priors more informative and further improve convergence.

Parameter $b$, denoting the amount of alcohol consumed during ‘binge’ drinking sessions, received an initial $b \sim\mathrm{Uniform}(5, 10)$ prior, later changed to the more informative $b \sim\mathrm{Normal}(6,1)$ prior, and finally to $b \sim\mathrm{Gamma}(4, 1.5)$ to anchor it away from $]-\infty, 0]$.

**Note:** No participant responded ‘Daily or almost daily’ in AUDIT-Q3, preventing from estimating the corresponding parameter. By default, we set $V_{5}= 5$.

The final model is reproduced in Code Block 1. Final priors and posteriors are plotted in Figure 2. Posterior parameters are summarised in Table 2 and sampling traces shown on Figure 4-Figure 6.

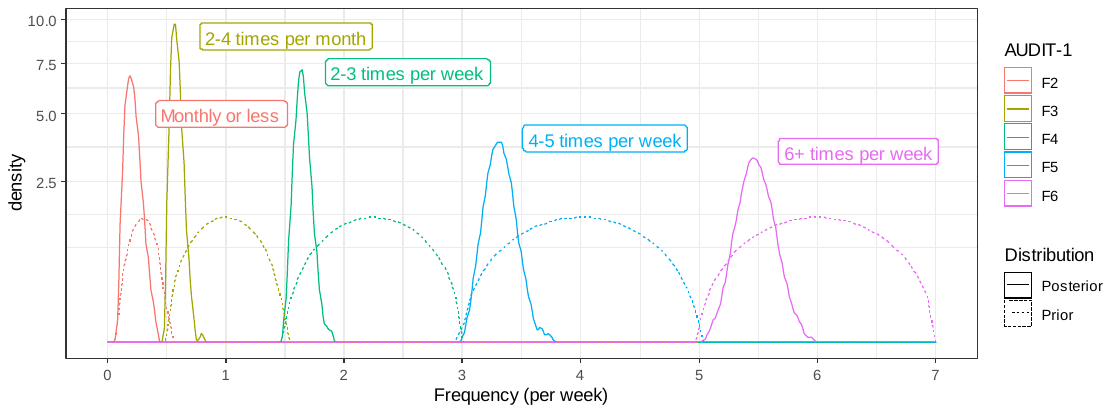

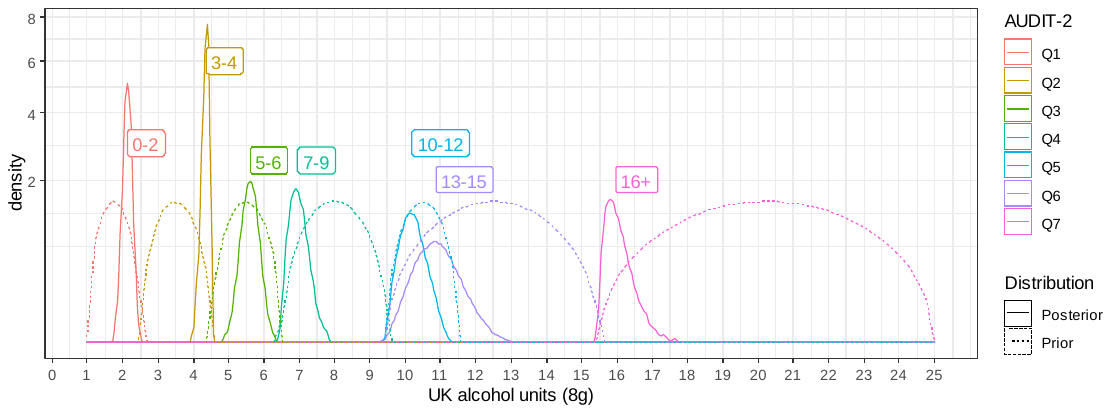

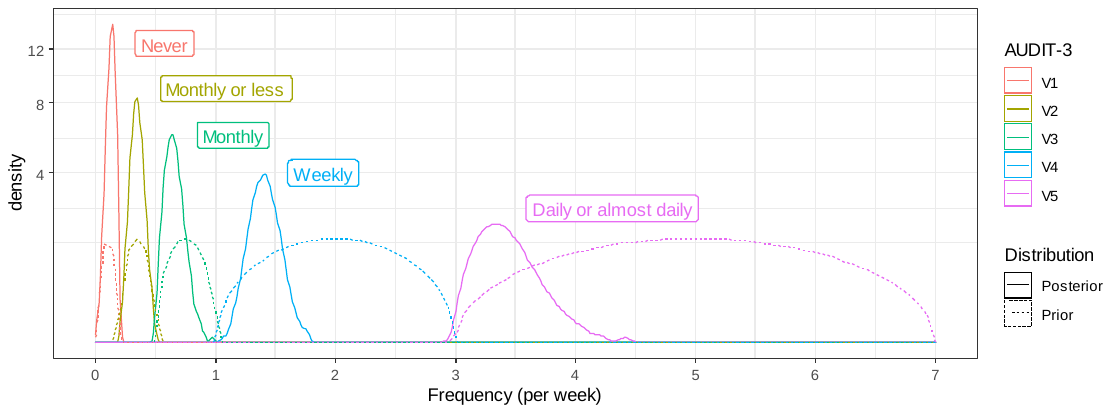

Figure 2 Kernel density plots of prior and posterior distributions of AUDIT response item values in the final model (after tuning)

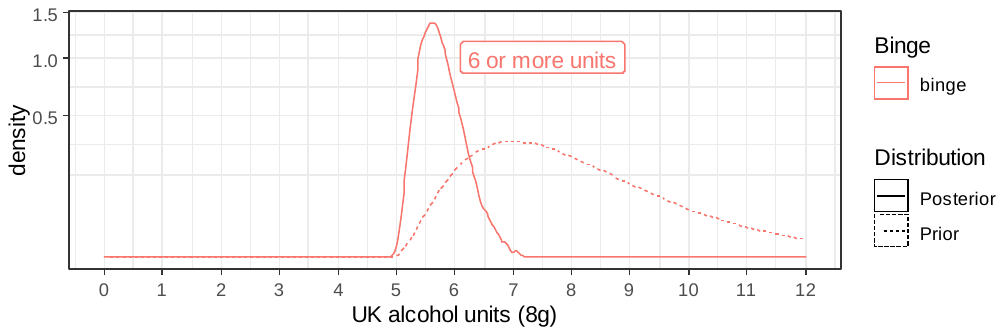

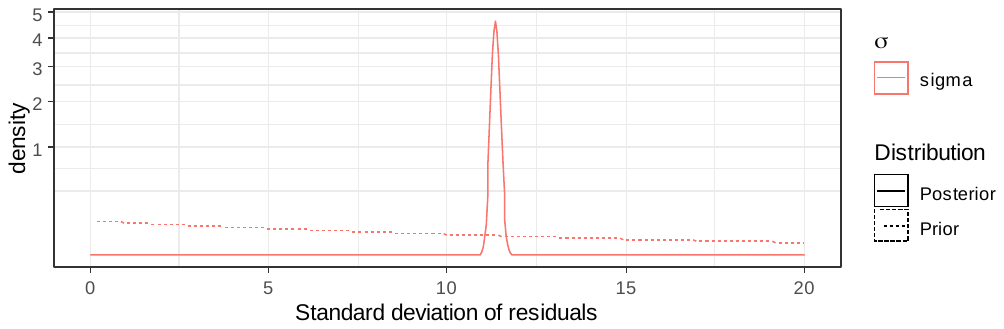

Figure 3 Kernel density plots of prior and posterior distributions of other model parameters in the final model (after tuning)

Table 2 Final posterior model parameters

|  |  |  |  | Percentiles | | | | |  |  |
| --- | --- | --- | --- | --- | --- | --- | --- | --- | --- | --- |
|  | Posterior  mean | Posterior  standard  error | Standard  deviation | 2.50% | 25% | 50% | 75% | 97.50% | ESS | Rhat |
| F2 | 0.21084 | 0.00083 | 0.05609 | 0.12094 | 0.16808 | 0.20525 | 0.24796 | 0.33126 | 4540 | 1.0002 |
| F3 | 0.57864 | 0.00061 | 0.03968 | 0.51510 | 0.54916 | 0.57455 | 0.60387 | 0.66579 | 4199 | 1.0001 |
| F4 | 1.64491 | 0.00090 | 0.05587 | 1.54400 | 1.60485 | 1.64304 | 1.68213 | 1.76120 | 3886 | 0.9999 |
| F5 | 3.31438 | 0.00165 | 0.09914 | 3.12223 | 3.24661 | 3.31337 | 3.38078 | 3.51141 | 3601 | 1.0002 |
| F6 | 5.47534 | 0.00236 | 0.12348 | 5.23515 | 5.39377 | 5.47317 | 5.55714 | 5.71733 | 2736 | 1.0007 |
| Q1 | 2.13199 | 0.00162 | 0.10225 | 1.93121 | 2.06266 | 2.13309 | 2.20096 | 2.33129 | 3981 | 1.0003 |
| Q2 | 4.35356 | 0.00159 | 0.08315 | 4.16115 | 4.30279 | 4.36570 | 4.41663 | 4.47803 | 2747 | 1.0004 |
| Q3 | 5.60338 | 0.00316 | 0.19533 | 5.21526 | 5.47115 | 5.60046 | 5.73547 | 5.98633 | 3809 | 1.0009 |
| Q4 | 6.95035 | 0.00312 | 0.20946 | 6.59599 | 6.79847 | 6.93031 | 7.08519 | 7.40503 | 4511 | 1.0014 |
| Q5 | 10.18389 | 0.00444 | 0.29359 | 9.66502 | 9.96738 | 10.16943 | 10.37890 | 10.79398 | 4367 | 1.0004 |
| Q6 | 10.85909 | 0.00715 | 0.50439 | 9.93277 | 10.50474 | 10.84133 | 11.19655 | 11.88402 | 4974 | 1.0001 |
| Q7 | 15.97190 | 0.00577 | 0.28838 | 15.56657 | 15.75252 | 15.92806 | 16.13885 | 16.67114 | 2500 | 1.0002 |
| V1 | 0.12909 | 0.00050 | 0.03202 | 0.06134 | 0.10833 | 0.13149 | 0.15251 | 0.18476 | 4138 | 1.0006 |
| V2 | 0.34295 | 0.00069 | 0.04791 | 0.25040 | 0.30965 | 0.34285 | 0.37586 | 0.43630 | 4831 | 1.0001 |
| V3 | 0.64481 | 0.00099 | 0.06493 | 0.53543 | 0.59649 | 0.63890 | 0.68701 | 0.78397 | 4300 | 1.0002 |
| V4 | 1.40232 | 0.00164 | 0.09945 | 1.21436 | 1.33444 | 1.40042 | 1.46857 | 1.60333 | 3693 | 1.0006 |
| binge | 5.70345 | 0.00487 | 0.28892 | 5.22719 | 5.49644 | 5.67047 | 5.88309 | 6.34626 | 3517 | 1.0000 |
| sigma | 11.35136 | 0.00124 | 0.10020 | 11.15421 | 11.28369 | 11.35164 | 11.41884 | 11.54630 | 6543 | 1.0001 |

ESS: effective sample size

Code block 1 Final STAN model syntax

```
data {
  int<lower=0> N;
  vector[N] GF;
  vector[N] audit1_2;
  vector[N] audit1_3;
  vector[N] audit1_4;
  vector[N] audit1_5;
  vector[N] audit1_6;
  vector[N] audit2_1;
  vector[N] audit2_2;
  vector[N] audit2_3;
  vector[N] audit2_4;
  vector[N] audit2_5;
  vector[N] audit2_6;
  vector[N] audit2_7;
  vector[N] audit3_1;
  vector[N] audit3_2;
  vector[N] audit3_3;
  vector[N] audit3_4;
  vector[N] audit3_5;
}

parameters {
  real<lower=0> sigma;
  real F2hyper;
  real F3hyper;
  real F4hyper;
  real F5hyper;
  real F6hyper;
  real Q1hyper;
  real Q2hyper;
  real Q3hyper;
  real Q4hyper;
  real Q5hyper;
  real Q6hyper;
  real Q7hyper;
  real V1hyper;
  real V2hyper;
  real V3hyper;
  real V4hyper;
  real V5hyper;
  real bingehyper;
}

transformed parameters{
  real F2 =  ((F2hyper * 0.4) + 0.1);
  real F3 =  ((F3hyper * 1) + 0.5);
  real F4 =  ((F4hyper * 1.5) + 1.5);
  real F5 =  ((F5hyper * 2) + 3);
  real F6 =  ((F6hyper * 2) + 5);
  real Q1 =  ((Q1hyper * 1.5) + 1);
  real Q2 =  ((Q2hyper * 2) + 2.5);
  real Q3 =  ((Q3hyper * 2) + 4.5);
  real Q4 =  ((Q4hyper * 3) + 6.5);
  real Q5 =  ((Q5hyper * 2) + 9.5);
  real Q6 =  ((Q6hyper * 6) + 9.5);
  real Q7 =  ((Q7hyper * 9.5) + 15.5);
  real V1 =  ((V1hyper * 0.2) + 0);
  real V2 =  ((V2hyper * 0.3) + 0.2);
  real V3 =  ((V3hyper * 0.5) + 0.5);
  real V4 =  ((V4hyper * 2) + 1);
  real V5 =  ((V5hyper * 4) + 3);
  real binge = bingehyper + 5;

  vector[N] mu;

  vector[N] F = (F2*audit1_2) + (F3*audit1_3) + (F4*audit1_4) + (F5*audit1_5) + (F6*audit1_6);
  vector[N] Q = (audit2_1 * Q1) + (audit2_2 * Q2) + (audit2_3 * Q3) + (audit2_4 * Q4) + (audit2_5 * Q5) + (audit2_6 * Q6) + (audit2_7 * Q7);
  vector[N] V = (audit3_1 * V1) + (audit3_2 * V2) + (audit3_3 * V3) + (audit3_4 * V4) + (audit3_5 * V5);

  mu =  (F .* Q) + (V * binge);
}

model {
  F2hyper ~ beta(2, 2);
  F3hyper ~ beta(2, 2);
  F4hyper ~ beta(2, 2);
  F5hyper ~ beta(2, 2);
  F6hyper ~ beta(2, 2);
  Q1hyper ~ beta(2, 2);
  Q2hyper ~ beta(2, 2);
  Q3hyper ~ beta(2, 2);
  Q4hyper ~ beta(2, 2);
  Q5hyper ~ beta(2, 2);
  Q6hyper ~ beta(2, 2);
  Q7hyper ~ beta(2, 2);
  V1hyper ~ beta(2, 2);
  V2hyper ~ beta(2, 2);
  V3hyper ~ beta(2, 2);
  V4hyper ~ beta(2, 2);
  V5hyper ~ beta(2, 2);

  bingehyper ~ gamma(4, 1.5);

  sigma ~ exponential(0.1);
  GF ~ normal(mu, sigma);
}
```

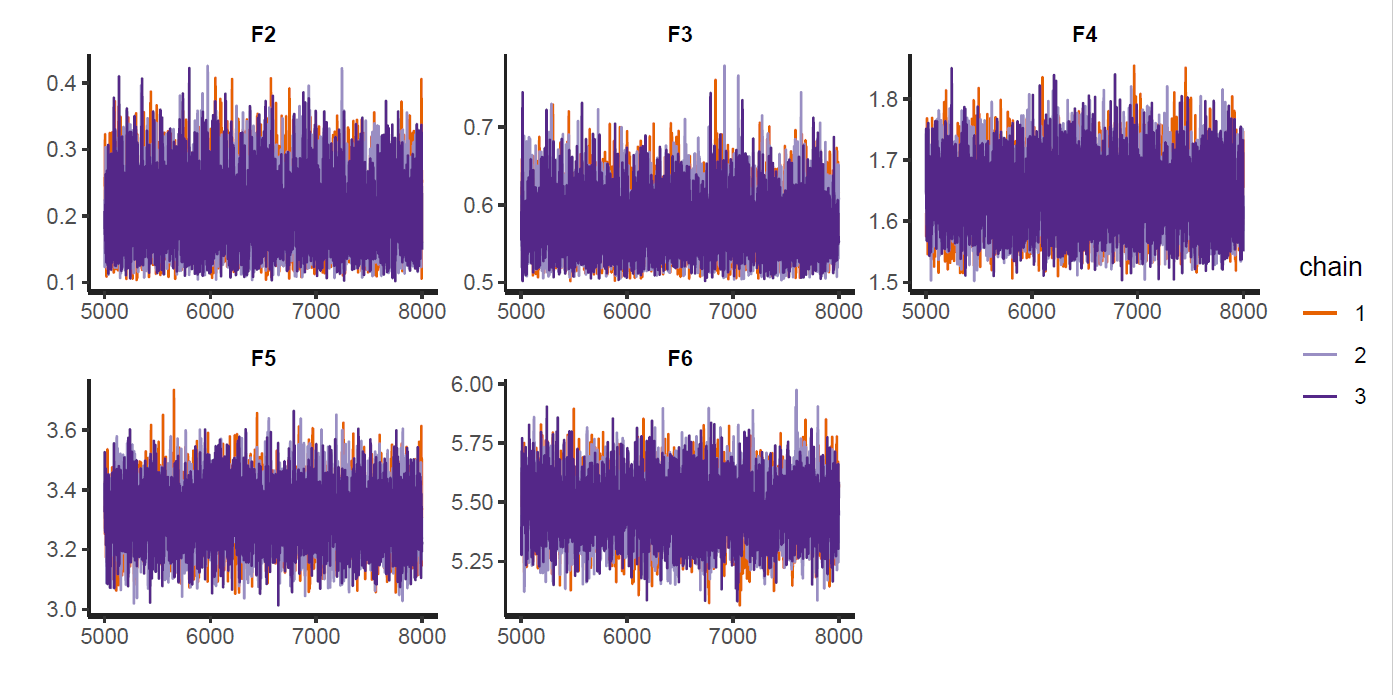

Figure 4 Hierarchical model estimation traces for the AUDIT-Q1 item (F2 = ‘Monthly or less’, …, F6 = ‘6 or more times a week’)

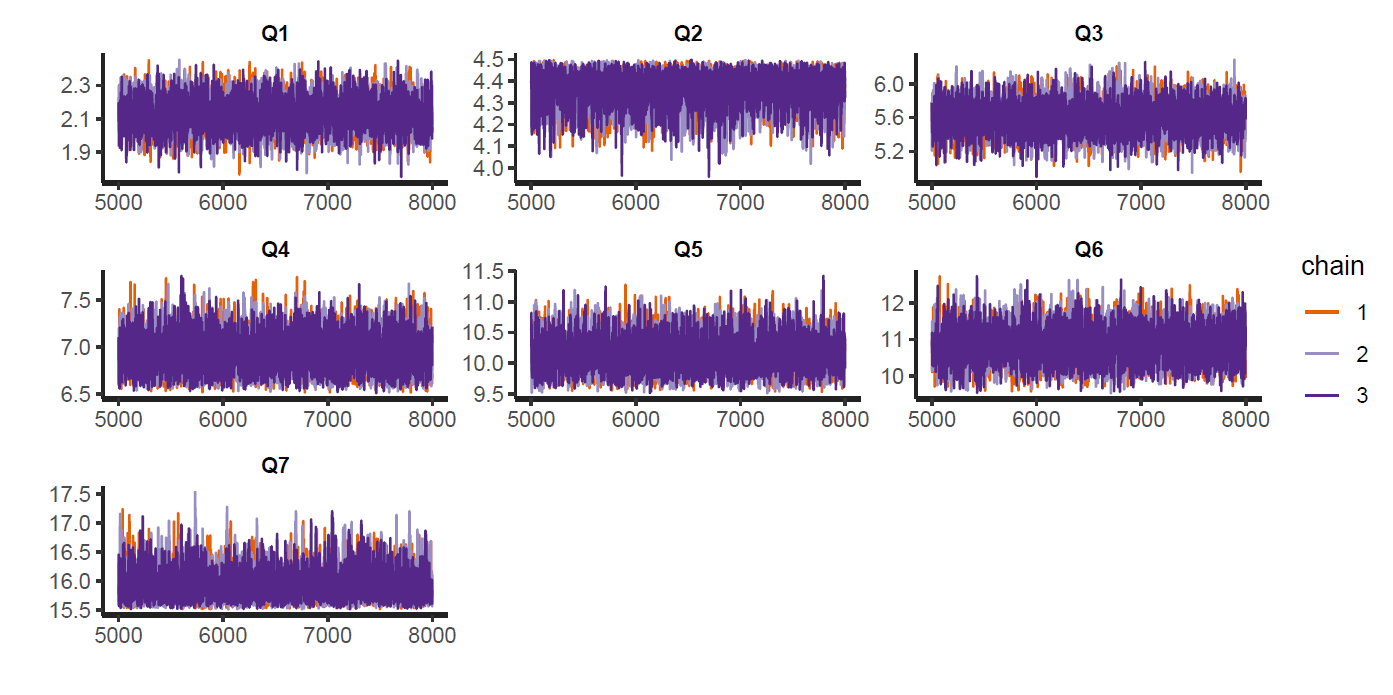

Figure 5 Hierarchical model estimation traces for the AUDIT-Q2 item (Q1 = ‘1 to 2’, …, Q7 = ’16 or more’)

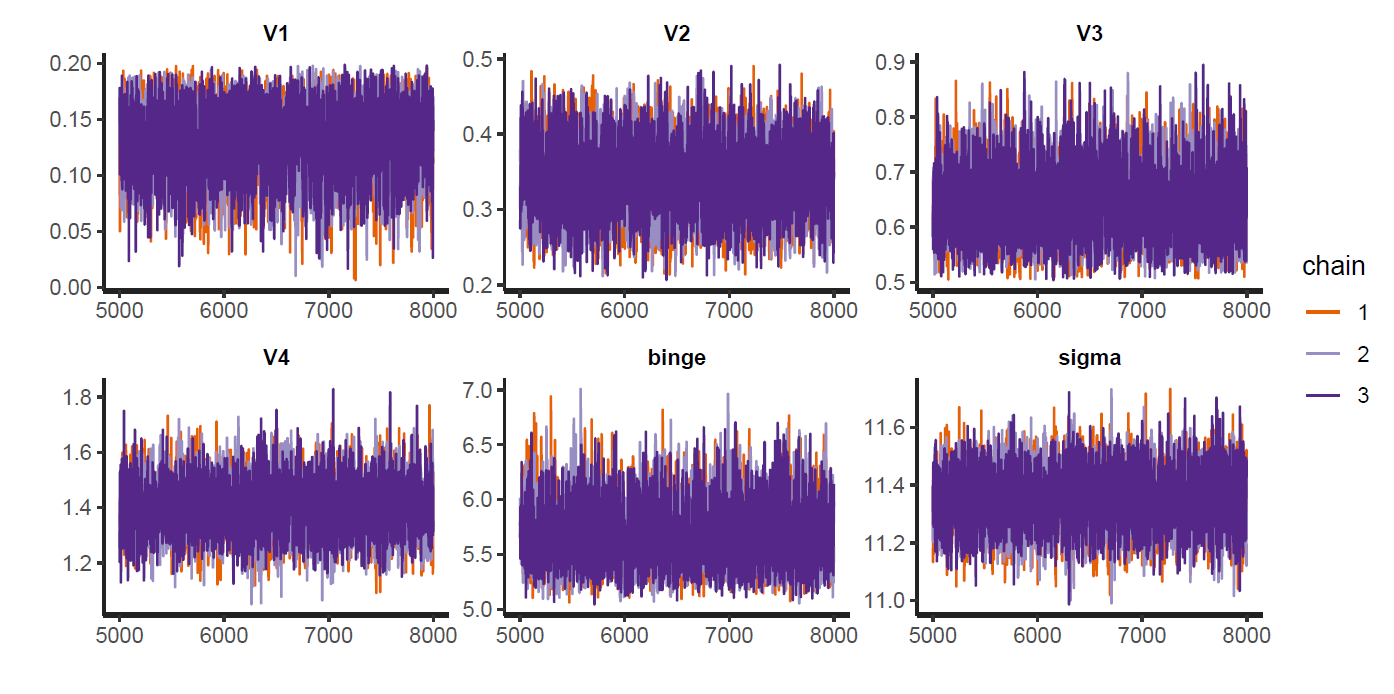

Figure 6 Hierarchical model estimation traces for the AUDIT-Q3 item (V1 = ‘Never’, …, V4 = ‘Weekly’)

### 4 Sensitivity analyses

Sensitivity analyses were conducted to identify the optimal method for computing the EWAC. Each of the four alternatives listed below was compared with the method reported in the paper. We found that each of them produced larger Mean Deviations and Root Mean Squared Deviations. They were thus discarded.

- AUDIT response item interval midpoint
- model-based coefficients estimated using random forest
- model-based coefficients estimated using STAN like in section 1, stratifying models by sex
- model-based coefficients estimated using STAN using a different estimating equation for $\mu$, based on the suggestion by Lemmens et al. [2] (Figure 7).

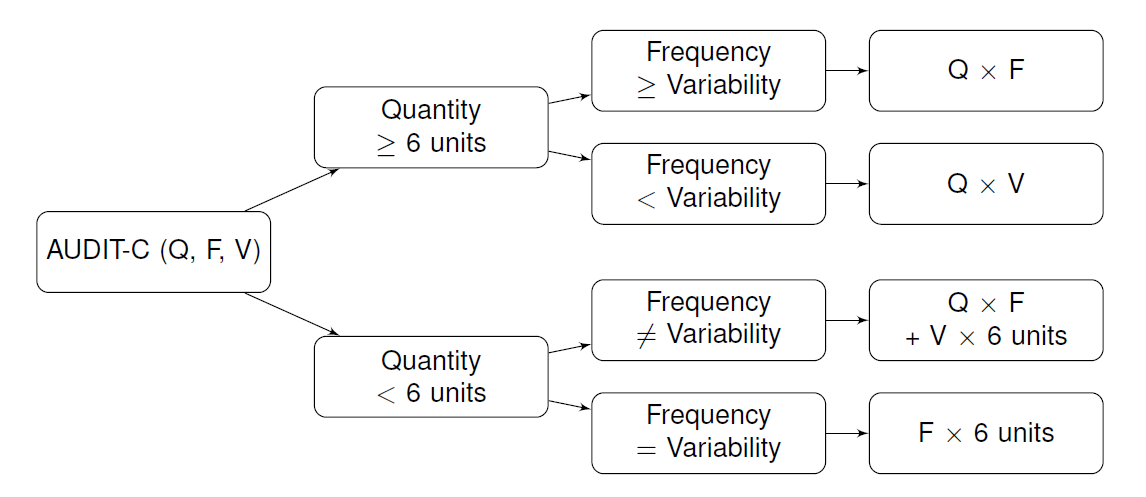

Figure 7 Quantity-Frequency-Variability estimator overview (adapted from Lemmens et al. [2])

### References

1.	Van Buuren S, Groothuis-Oudshoorn K. Multivariate Imputation by Chained Equations. *J Stat Softw*. PMID:22289957

2.	Lemmens P et al. Measuring quantity and frequency of drinking in a general population survey: a comparison of five indices. *J Stud Alcohol*. PMID:1405641
