## Supplementary material for "Concurrent validity of an Estimator of Weekly Alcohol Consumption (EWAC) based on the Extended AUDIT": S2: ATS AUDIT questionnaire

### **Supplementary information S2: Alcohol Toolkit Study Extended AUDIT questionnaire schedule**

The next few questions form part of a study about consumption of alcohol. We understand that this is a highly sensitive topic and would therefore like to remind you that any information you give me is strictly confidential and will be used for research purposes only. Some questions asked may not necessarily apply to you.

**NEW SCREEN**

These first few questions ask about the alcohol you have drunk **in the last 6 months**, including about how many standard drinks you have consumed. Please note that 1 standard drink equals 1 unit of alcohol. So, for example, a pint of regular beer or lager is equal to 2 standard drinks or 2 units. A description of a standard drink is given in the show prompt here.

Please be aware that all your answers will be handled confidentially.

**INTERVIEWER: HAND SHOW PROMPT TA TO RESPONDENT FOR REMAINDER OF MODULE**

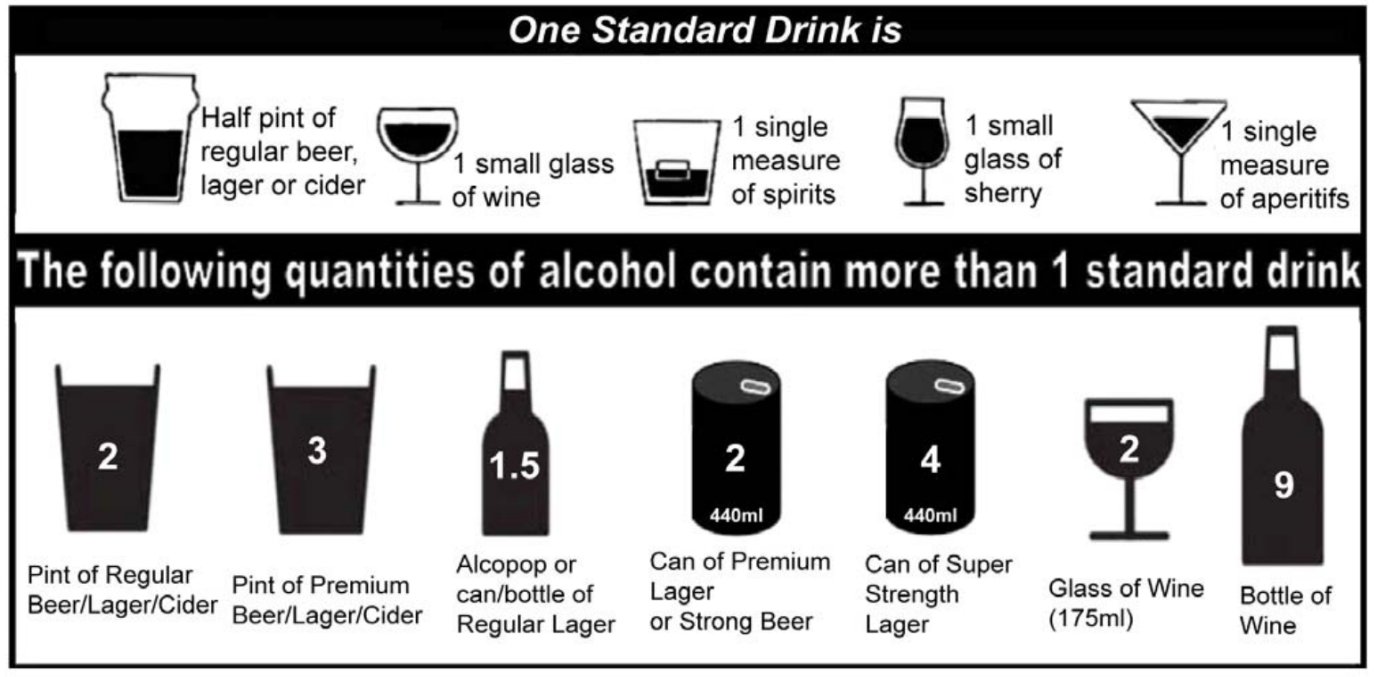

**.showscreen**

**ASK ALL**

**audit1.** How often do you have a drink containing alcohol?

**SP, ALLOW DK, REF**

0. Never

1. Monthly or less

2. 2 to 4 times a month

3. 2 to 3 times a week

4. 4 to 5 times a week

5. 6 or more times a week

**[skip to audit9 if audit1=0]**

**ASK ALL EXCEPT 0 AT audit1**

**audit2.** How many standard drinks containing alcohol do you have on a typical day when you are drinking?

Please work this out based on the ‘standard drink’ definitions included on the show prompt.

**INTERVIEWER: IF RESPONDENT SAYS ‘DON’T KNOW’ ENCOURAGE THEM TO GIVE BEST ESTIMATE**

**SP, ALLOW DK, REF**

0. 1 to 2

1. 3 to 4

2. 5 to 6

3. 7 to 9

4. 10 to 12

5. 13 to 15

6. 16 or more

**ASK ALL EXCEPT 0 AT audit1**

**audit3.** How often do you have six or more standard drinks on one occasion?

If you need to remind yourself of the definition of a ‘standard drink’, please see the definitions on the show prompt.

**SP, ALLOW DK, REF**

0. Never

1 Less than monthly

2 Monthly

3 Weekly

4 Daily or almost daily

**[Skip to audit9 if audit2 and audit3 are both 0.]**

**ASK ALL EXCEPT THOSE ANSWERING CODE 0 AT audit2 AND audit3**

**audit4.** How often during the last 6 months have you found that you were not able to stop drinking once you had started?

**SP, ALLOW DK, REF**

0. Never

1 Less than monthly

2 Monthly

3 Weekly

4 Daily or almost daily

**ASK ALL EXCEPT THOSE ANSWERING CODE 0 AT audit2 AND audit3**

**audit5.** How often during the last 6 months have you failed to do what was normally expected from you because of drinking?

**SP, ALLOW DK, REF**

0. Never

1 Less than monthly

2 Monthly

3 Weekly

4 Daily or almost daily

**ASK ALL EXCEPT THOSE ANSWERING CODE 0 AT audit2 AND audit3**

**audit6.** How often during the last 6 months have you needed a first drink in the morning to get yourself going after a heavy drinking session?

**SP, ALLOW REF, DK**

0. Never

1 Less than monthly

2 Monthly

3 Weekly

4 Daily or almost daily

**ASK ALL EXCEPT THOSE ANSWERING CODE 0 AT audit2 AND audit3**

**audit7.** How often during the last 6 months have you had a feeling of guilt or remorse after drinking?

**SP, ALLOW REF, DK**

0. Never

1 Less than monthly

2 Monthly

3 Weekly

4 Daily or almost daily

**ASK ALL EXCEPT THOSE ANSWERING CODE 0 AT audit2 AND audit3**

**audit8.** How often during the last 6 months have you been unable to remember what happened the night before because you had been drinking?

**SP, ALLOW REF, DK**

0. Never

1 Less than monthly

2 Monthly

3 Weekly

4 Daily or almost daily

**ASK ALL**

**audit9.** Have you or someone else ever been injured as a result of your drinking?

**SP, ALLOW REF, DK**

0. No

1. Yes, but not in the last 6 months

2. Yes, during the last 6 months

**ASK ALL**

**audit10.** Has a relative or friend or a doctor or another health worker ever been concerned about your drinking or suggested you cut down?

**SP, ALLOW REF, DK**

0. No

1. Yes, but not in the last 6 months

2. Yes, during the last 6 months
