## Supplementary material for "Concurrent validity of an Estimator of Weekly Alcohol Consumption (EWAC) based on the Extended AUDIT": S3: ATS Graduated Frequency questionnaire

### **Supplementary information S3: Alcohol Toolkit Study Graduated-Frequency questionnaire schedule**

**ASK IF CODES 2-6 AT audit1.**

**TA47_01.** On how many days, if any, did you personally drink a drink containing alcohol in the last four weeks?

**NUMERIC, ALLOW ANSWER 0-28, ALLOW DK & REF**

**ASK IF >0 AT TA47_01.**

**TA47_02.**

As you may be aware, the amount of alcohol in contained in a drink is measured in units.

What was the maximum number of units you personally consumed on any one day when drinking an alcoholic drink or drinks in the last four weeks?

If you need to remind yourself of the definition of a ‘standard drink’ or ‘unit’, please see the definitions on the show prompt.

**INTERVIEWER: SHOW TA SHOWCARD**

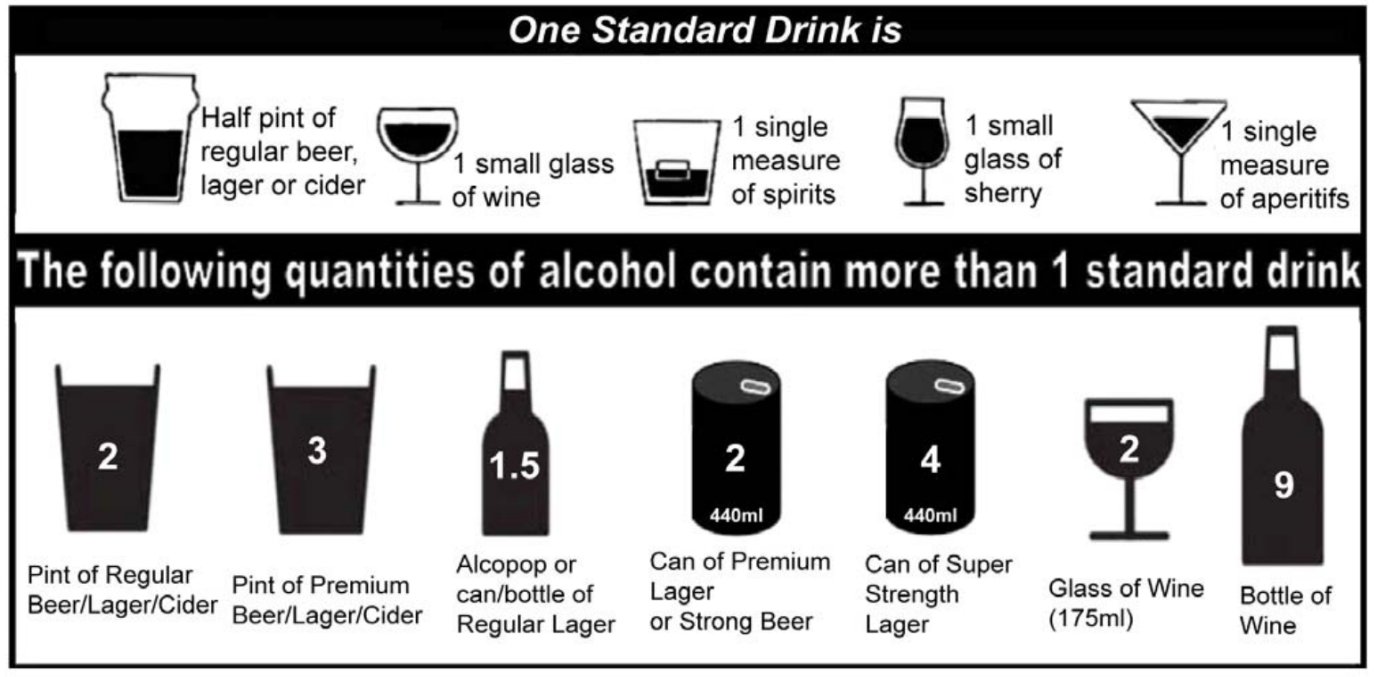

**NUMERIC, ALLOW ANSWER 1-60, ALLOW DK & REF**

**ASK IF (>=1 AT TA47_01) AND (1-60 AT TA47_02)**

**TA47_03.**

You mentioned that in the last four weeks you personally had a drink containing alcohol on **[INSERT ANSWER FROM TA47_01]** days.

On how many days, if any, in the last four weeks did you personally drink…

**INTERVIEWER: IF NECESSARY, SHOW SHOWCARD TA**

**NUMERIC ANSWER PER STATEMENT, ALLOW DK AND REF**

**SUM OF ANSWERS ACROSS STATEMENTS MUST EQUAL ANSWER AT TA47_01**

**FOR CODES 2-9, IF TA47_02 ANSWER FALLS WITHIN THESE BANDS, SHOW UPPER LIMIT FOR THESE BANDS AS TA47_02 MINUS 1.**

1. **[INSERT ANSWER FROM TA47_02]** units? **SHOW TO ALL QUALIFYING – ANSWER MUST BE >=1**

2. 51-60 units? **ASK IF >51 AT TA47_02**

3. 41-50 units? **ASK IF >41 AT TA47_02**

4. 31-40 units? **ASK IF >31 AT TA47_02**

5. 21-30 units? **ASK IF >21 AT TA47_02**

6. 16-20 units? **ASK IF >16 AT TA47_02**

7. 11-15 units? **ASK IF >11 AT TA47_02**

8. 8-10 units? **ASK IF >8 AT TA47_02**

9. 5-7 units? **ASK IF >5 AT TA47_02**

10. 3-4 units? **ASK IF >3 AT TA47_02**

11. 1-2 units? **ASK IF >1 AT TA47_02**
