## Supplementary material for "Concurrent validity of an Estimator of Weekly Alcohol Consumption (EWAC) based on the Extended AUDIT": S5: Subgroup analyses

### Supplementary information S5: Analysis of subgroup variation in bias and error

MDs and RMSDs were regressed against respondent characteristics to identify potential subgroup differences in bias or precision from the reference category (White British males aged 25–34 years without educational qualifications, who never smoked). Model predictors accounted for <5% of the variance, indicating stability in MD and RMSD across subgroups (Tables S5.1-S5.2).

Figure S5.1 shows subgroups with a predicted MD exceeding $\pm$ 1 unit/week and a coefficient $p$-value<0.05. Respondents of Black/Other/White Other ethnic groups had an overestimated EWAC: their MDs exceeded the reference MD by 1.7 units [0.4, 2.9]; 2.3 units [0.1, 4.4]; and 1.0 units [0.3, 1.7] respectively. MDs of men aged 55–64 years, or $\geq$ 75 years exceeded the reference MD by 1.1 units [0.2, 2.0] and 1.0 units [0.02, 2.0] respectively. Recent attempts to reduce alcohol intake in increasing-risk drinkers were associated with a modest underestimation in EWAC by 0.9 UK units [0.1, 1.6] (Table S5.2).

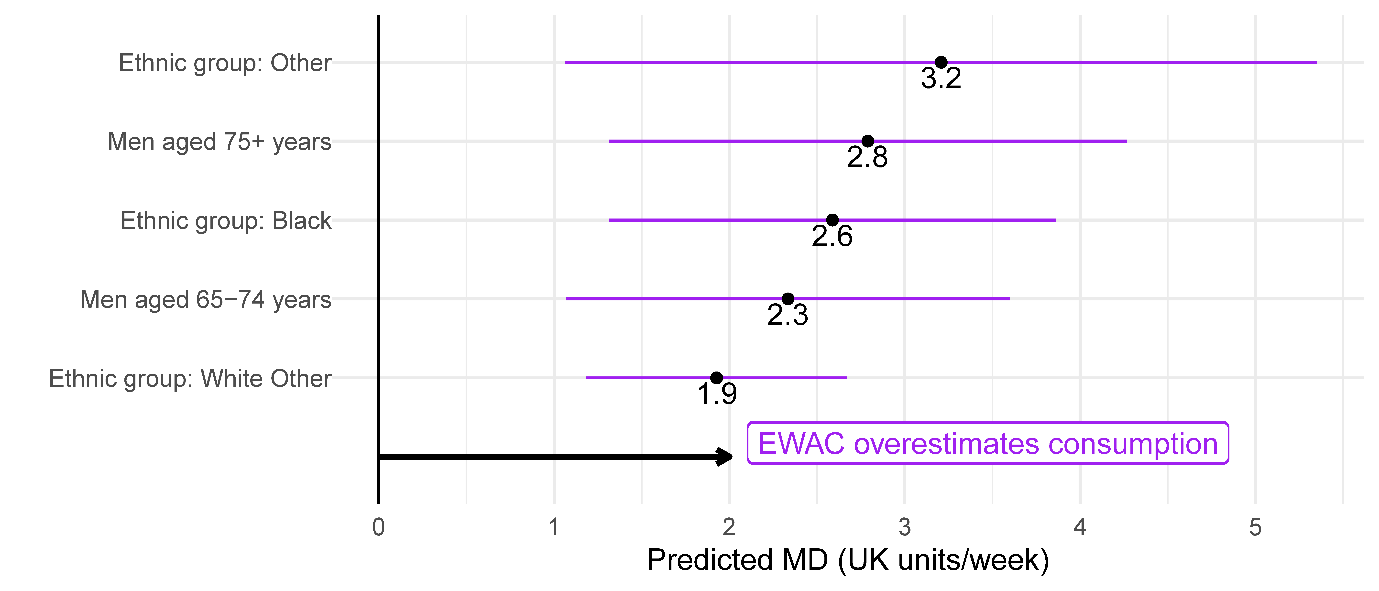

*Figure S5.1:* Forest plot of modelled MD for selected subgroups

Figure S5.2 shows subgroups with an RMSD at least 25% smaller/greater than the reference. RMSD was 53% [44%, 63%] greater in current smokers and 45% [21%, 75%] greater in respondents who stopped smoking in the past year. It was also 43% [27%, 62%] and 41% [24%, 60%] greater respectively in men and respondents aged 16–24 years. In contrast, error was 20 to 40% smaller in Black/Asian/Other/White Other ethnic groups.

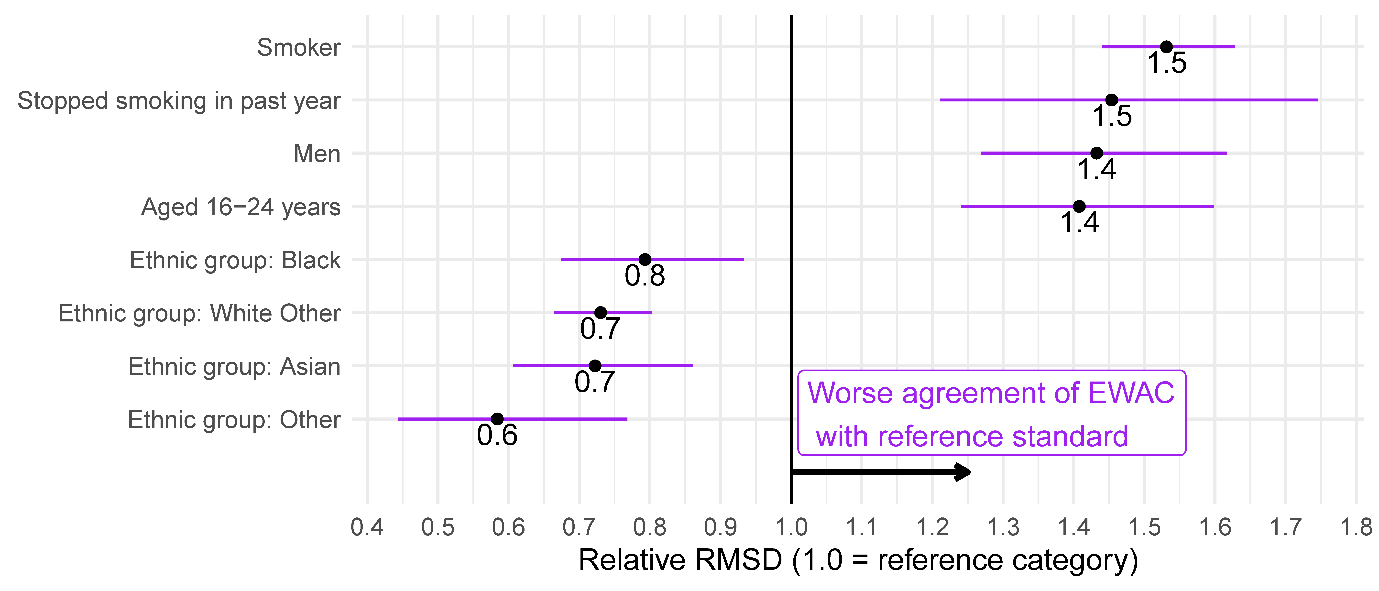
 *Figure S5.2:* Forest plot of RMSD ratio (selected subgroups to reference category)

Figure S5.3 presents the same metrics for increasing-/higher-risk drinkers (AUDIT-C $\geq$ 5 or AUDIT $\geq$ 8). Educational qualifications significantly improved the agreement between EWAC and the reference standard in this group (Table S5.2). School and degree-level qualifications reduced RMSD by 22% [12%, 31%] and 28% [19%, 37%] respectively, suggesting that respondents may have better recall and clarity over alcohol beverage content.

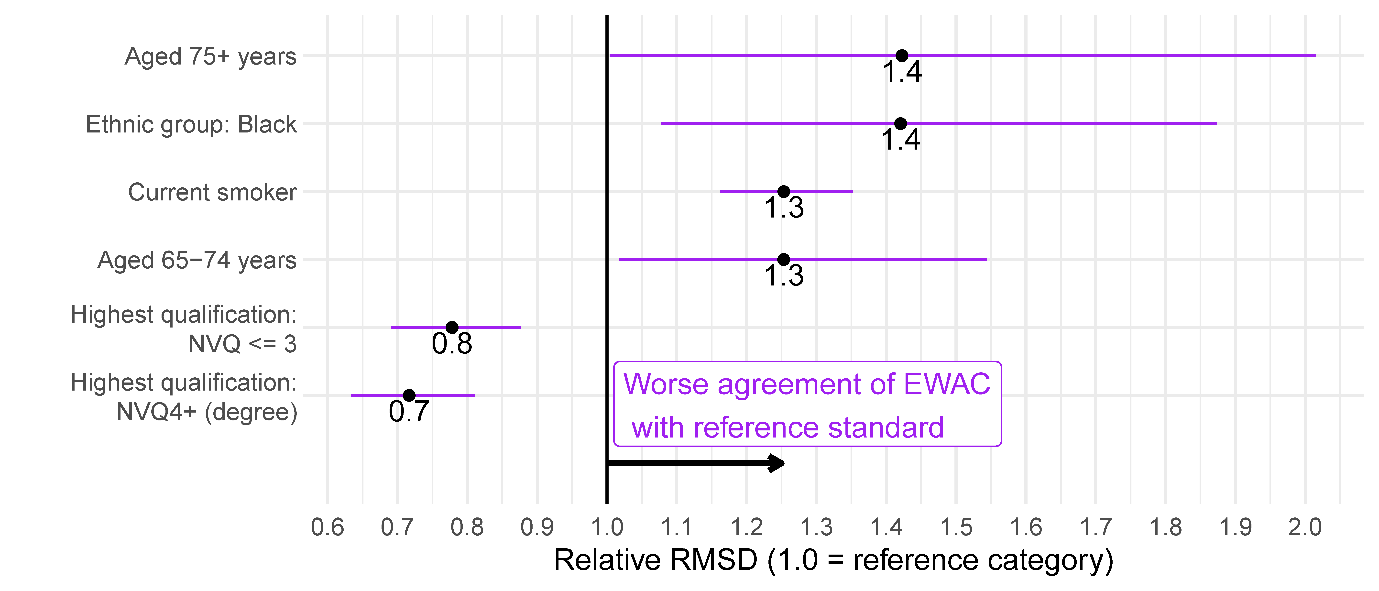

*Figure S5.3:* Forest plot of RMSD ratio (selected subgroups to reference category) in respondents with a hazardous/harmful alcohol use (AUDIT-C>=5 or AUDIT>=8)

**Table S5.1:** Coefficients of linear regression of subgroup bias and error of EWAC compared with GF in all respondents (n = 15,373)

|  | EWAC - GF  Coefficient (95% CI) | log[ (EWAC-GF)^2^ ]  Coefficient (95% CI) |
| --- | --- | --- |
| Intercept | 0.9 (-0.01, 1.9) | 0.7 (0.5, 1.0)*** |
| Male | -1.1 (-2.0, -0.1)* | 0.7 (0.5, 1.0)*** |
| Age group |  |  |
| 16-24 years | 0.9 (-0.1, 1.9) | 0.7 (0.4, 0.9)*** |
| 25-34 years | 0.0 (*reference*) | 0.0 (*reference*) |
| 35-44 years | -0.4 (-1.3, 0.6) | 0.1 (-0.2, 0.3) |
| 45-54 years | -0.2 (-1.1, 0.7) | 0.2 (-0.03, 0.4) |
| 55-64 years | 0.4 (-0.5, 1.3) | 0.1 (-0.1, 0.4) |
| 65-74 years | -0.3 (-1.2, 0.7) | -0.2 (-0.5, 0.02) |
| 75+ years | -0.8 (-2.0, 0.3) | -0.2 (-0.5, 0.1) |
| Male × age interactions |  |  |
| Male × 16-24 years | -0.5 (-1.9, 0.8) | -0.2 (-0.5, 0.1) |
| Male × 35-44 years | -0.5 (-1.8, 0.8) | -0.04 (-0.4, 0.3) |
| Male × 45-54 years | -0.7 (-2.0, 0.6) | 0.1 (-0.2, 0.4) |
| Male × 55-64 years | -0.1 (-1.4, 1.1) | 0.1 (-0.2, 0.5) |
| Male × 65-74 years | 1.4 (0.1, 2.7)* | 0.3 (0.02, 0.7)* |
| Male × 75+ years | 1.9 (0.4, 3.3)* | -0.3 (-0.7, 0.1) |
| Ethnic group |  |  |
| White British | 0.0 (*reference*) | 0.0 (*reference*) |
| White Other | 1.0 (0.3, 1.7)** | -0.6 (-0.8, -0.4)*** |
| Mixed | -0.5 (-2.0, 0.9) | -0.3 (-0.7, 0.1) |
| Asian | 1.3 (-0.1, 2.7) | -0.7 (-1.0, -0.3)*** |
| Black | 1.7 (0.4, 2.9)* | -0.5 (-0.8, -0.1)** |
| Other | 2.3 (0.1, 4.4)* | -1.1 (-1.6, -0.5)*** |
| Religion |  |  |
| No religion | 0.0 (*reference*) | 0.0 (*reference*) |
| Christian | 0.04 (-0.3, 0.4) | -0.2 (-0.3, -0.1)*** |
| Muslim | 0.5 (-2.4, 3.5) | -0.5 (-1.3, 0.2) |
| Any other religion | -0.7 (-1.7, 0.3) | -0.1 (-0.3, 0.2) |
| Highest qualification |  |  |
| No qualification | 0.0 (*reference*) | 0.0 (*reference*) |
| NVQ < = 3 | -0.1 (-0.8, 0.5) | 0.02 (-0.1, 0.2) |
| NVQ4+ (degree) | -0.2 (-0.9, 0.4) | 0.1 (-0.1, 0.3) |
| Other | -0.1 (-0.9, 0.8) | 0.1 (-0.1, 0.3) |
| Smoking status |  |  |
| Never smoker | 0.0 (*reference*) | 0.0 (*reference*) |
| Stopped> 1y ago | -0.1 (-0.5, 0.4) | 0.4 (0.3, 0.5)*** |
| Stopped in past year | -0.6 (-2.1, 0.8) | 0.7 (0.4, 1.1)*** |
| Current smoker | -0.7 (-1.2, -0.2)** | 0.9 (0.7, 1.0)*** |
| Observations | 15,373 | 15,373 |
| R^2^ | 0.01 | 0.05 |
| Residual Std. Error (df = 15345) | 10.7 | 2.7 |
| *F* Statistic (df = 27; 15345) | 4.1*** | 29.2*** |

Notes:	**p*<0.05; ***p*<0.01; ****p*<0.001

**Table S5.2:** Coefficients of linear regression of the subgroup bias and error of EWAC compared with GF in respondents with a hazardous/harmful alcohol use (AUDIT-C>=5 or AUDIT>=8; n = 6,909)

|  | EWAC - GF  Coefficient (95% CI) | log[ (EWAC-GF)^2^ ]  Coefficient (95% CI) |
| --- | --- | --- |
| Intercept | 0.5 (-1.6, 2.5) | 0.5 (-1.6, 2.5) |
| Male | -1.1 (-3.0, 0.8) | -1.1 (-3.0, 0.8) |
| Age group |  |  |
| 16-24 years | 1.1 (-0.8, 3.0) | 1.1 (-0.8, 3.0) |
| 25-34 years | 0.0 (*reference*) | 0.0 (*reference*) |
| 35-44 years | -0.4 (-2.4, 1.6) | -0.4 (-2.4, 1.6) |
| 45-54 years | 0.3 (-1.6, 2.2) | 0.3 (-1.6, 2.2) |
| 55-64 years | 2.6 (0.6, 4.6)* | 2.6 (0.6, 4.6)* |
| 65-74 years | 1.1 (-1.2, 3.5) | 1.1 (-1.2, 3.5) |
| 75+ years | 5.2 (1.2, 9.1)* | 5.2 (1.2, 9.1)* |
| Male × age interactions |  |  |
| Male × 16-24 years | -0.9 (-3.3, 1.6) | -0.9 (-3.3, 1.6) |
| Male × 35-44 years | -1.4 (-4.0, 1.1) | -1.4 (-4.0, 1.1) |
| Male × 45-54 years | -1.4 (-3.8, 1.1) | -1.4 (-3.8, 1.1) |
| Male × 55-64 years | -1.6 (-4.1, 0.9) | -1.6 (-4.1, 0.9) |
| Male × 65-74 years | 1.1 (-1.7, 3.9) | 1.1 (-1.7, 3.9) |
| Male × 75+ years | -2.3 (-6.7, 2.2) | -2.3 (-6.7, 2.2) |
| Ethnic group |  |  |
| White British | 0.0 (*reference*) | 0.0 (*reference*) |
| White Other | 1.6 (-0.01, 3.2) | 1.6 (-0.01, 3.2) |
| Mixed | -1.4 (-4.3, 1.4) | -1.4 (-4.3, 1.4) |
| Asian | 2.8 (-0.6, 6.3) | 2.8 (-0.6, 6.3) |
| Black | 4.2 (1.1, 7.3)** | 4.2 (1.1, 7.3)** |
| Other | 5.4 (0.3, 10.5)* | 5.4 (0.3, 10.5)* |
| Religion |  |  |
| No religion | 0.0 (*reference*) | 0.0 (*reference*) |
| Christian | 0.2 (-0.5, 0.9) | 0.2 (-0.5, 0.9) |
| Muslim | 2.2 (-8.6, 13.1) | 2.2 (-8.6, 13.1) |
| Any other religion | -2.5 (-4.5, -0.4)* | -2.5 (-4.5, -0.4)* |
| Highest qualification |  |  |
| No qualification | 0.0 (*reference*) | 0.0 (*reference*) |
| NVQ < = 3 | 0.7 (-0.7, 2.0) | 0.7 (-0.7, 2.0) |
| NVQ4+ (degree) | 0.5 (-0.9, 1.9) | 0.5 (-0.9, 1.9) |
| Other | 0.1 (-1.7, 1.9) | 0.1 (-1.7, 1.9) |
| Smoking status |  |  |
| Never smoker | 0.0 (*reference*) | 0.0 (*reference*) |
| Stopped> 1y ago | -0.5 (-1.4, 0.3) | -0.5 (-1.4, 0.3) |
| Stopped in past year | -1.0 (-3.5, 1.4) | -1.0 (-3.5, 1.4) |
| Current smoker | -1.4 (-2.3, -0.6)** | -1.4 (-2.3, -0.6)** |
| Favourite drink |  |  |
| Beer | 0.0 (*reference*) | 0.0 (*reference*) |
| Cider | -0.6 (-2.2, 1.0) | -0.6 (-2.2, 1.0) |
| Mixed spirits | 1.2 (-0.3, 2.7) | 1.2 (-0.3, 2.7) |
| Other | 4.5 (0.4, 8.6)* | 4.5 (0.4, 8.6)* |
| Spirits alone | 1.2 (-0.1, 2.6) | 1.2 (-0.1, 2.6) |
| Wine | 0.5 (-0.4, 1.4) | 0.5 (-0.4, 1.4) |
| Attempts to cut down |  |  |
| No attempt to cut down | 0.0 (*reference*) | 0.0 (*reference*) |
| Attempt to cut down in last 12 months | -0.9 (-1.6, -0.1)* | -0.9 (-1.6, -0.1)* |
| Observations | 6,909 | 6,909 |
| R^2^ | 0.02 | 0.02 |
| Residual Std. Error (df = 9816) | 14.1 | 14.1 |
| *F* Statistic (df = 33; 9816) | 4.3^***^ | 4.3^***^ |

Notes:	**p*<0.05; ***p*<0.01; ****p*<0.001
