## Supplementary material for "Concurrent validity of an Estimator of Weekly Alcohol Consumption (EWAC) based on the Extended AUDIT": S6: ROC curves

### Supplementary Figure S6: Receiver operating characteristics curves of the AUDIT-C, full AUDIT, and EWAC on consumption thresholds from the GF reference standard

Note: The raw data for every ROC curve are available from the repository <https://github.com/peterdutey/ewac-validation/tree/master/04_suppmat/ROC>

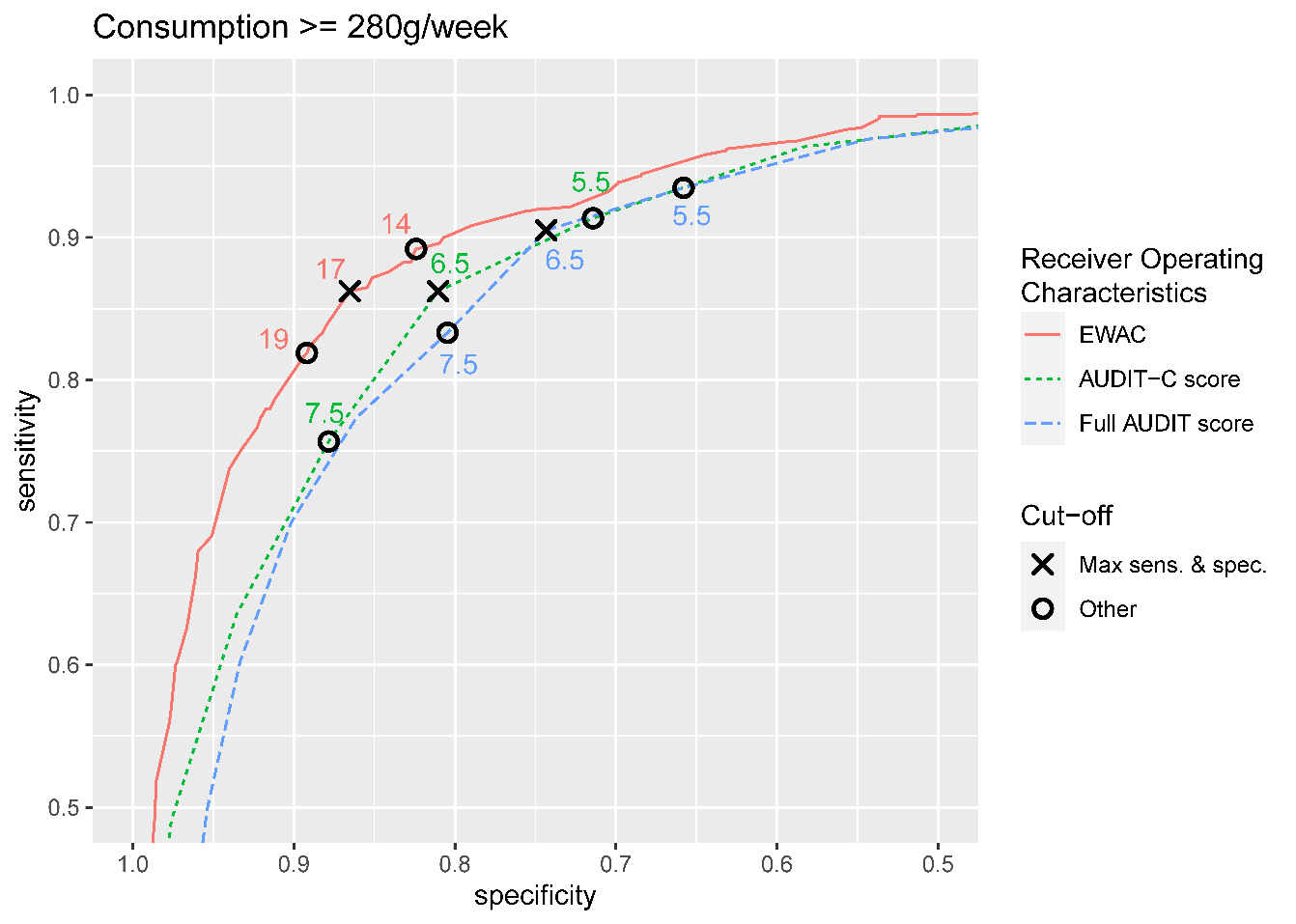

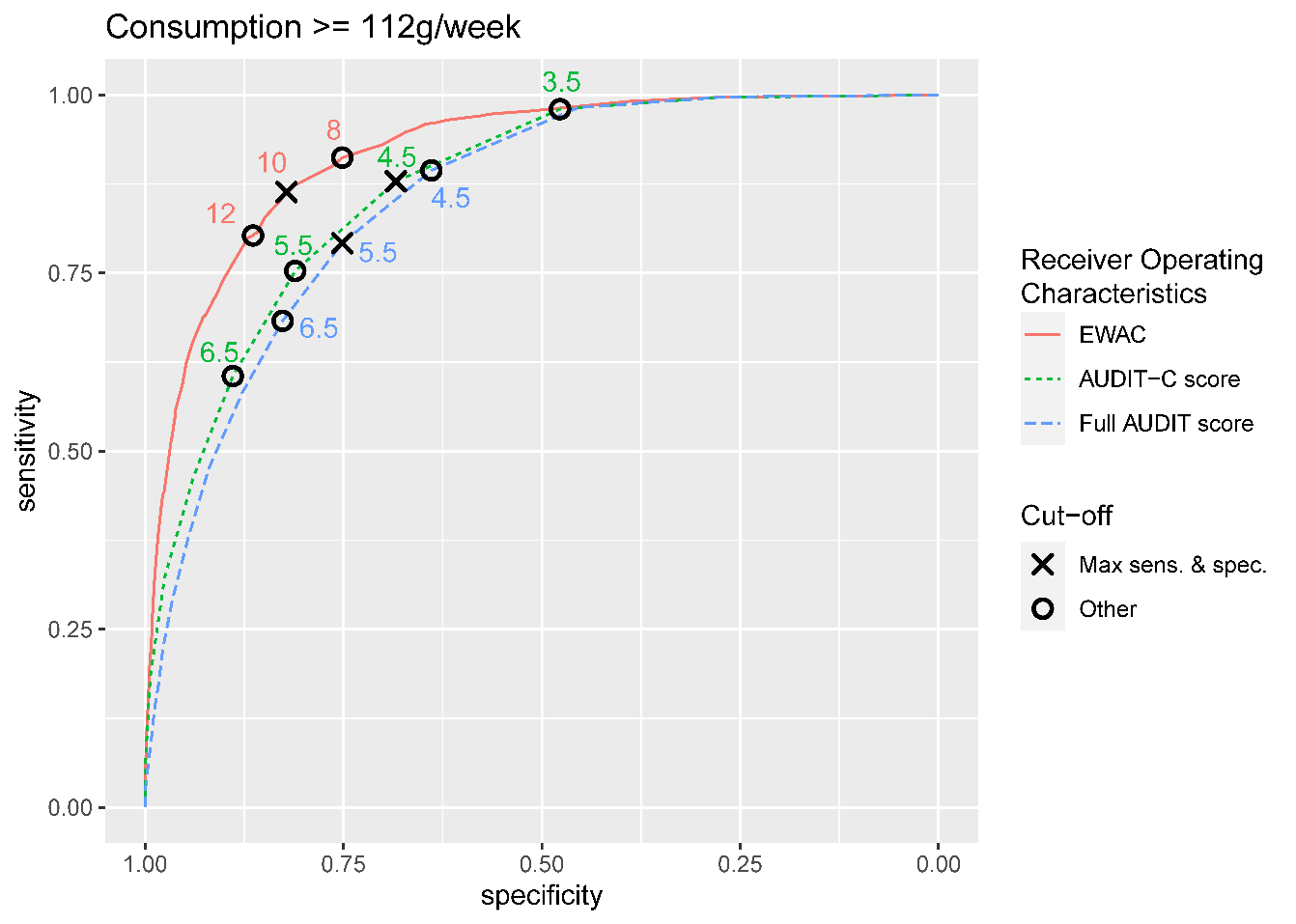
