## Supplementary material for "Concurrent validity of an Estimator of Weekly Alcohol Consumption (EWAC) based on the Extended AUDIT": S7: Demographics

### Supplementary Table S7: Demographic characteristics and alcohol consumption

|  | **Alcohol consumption in units/week (Graduated Frequency)** | | | | | |  |
| --- | --- | --- | --- | --- | --- | --- | --- |
|  | **Never drinks** (AUDIT-1=0)  (*N=*14399) | **<14**  (*N=*17402) | **14-34**  (*N=*3932) | **35-50**  (*N=*582) | **50-400**  (*N=*483) | *Missing*  (*N=*4017) | **Total** (*N=*40815) |
|  | n (%) | n (%) | n (%) | n (%) | n (%) | n (%) | N (%) |
| **Gender** |  |  |  |  |  |  |  |
| Women | 7996 (55.5%) | 8713 (50.1%) | 1200 (30.5%) | 124 (21.3%) | 104 (21.5%) | 2200 (54.8%) | 20337 (49.8%) |
| Men | 6403 (44.5%) | 8689 (49.9%) | 2732 (69.5%) | 458 (78.7%) | 379 (78.5%) | 1817 (45.2%) | 20478 (50.2%) |
| **Age** |  |  |  |  |  |  |  |
| 16-24 years | 2383 (16.7%) | 2246 (13.0%) | 491 (12.5%) | 72 (12.4%) | 56 (11.6%) | 671 (16.9%) | 5919 (14.6%) |
| 25-34 years | 2400 (16.8%) | 2320 (13.4%) | 394 (10.0%) | 57 (9.8%) | 43 (8.9%) | 582 (14.7%) | 5796 (14.3%) |
| 35-44 years | 2243 (15.7%) | 2378 (13.7%) | 463 (11.8%) | 57 (9.8%) | 71 (14.8%) | 514 (13.0%) | 5726 (14.1%) |
| 45-54 years | 1845 (13.0%) | 2852 (16.5%) | 678 (17.3%) | 124 (21.3%) | 95 (19.8%) | 529 (13.3%) | 6123 (15.1%) |
| 55-64 years | 1808 (12.7%) | 2946 (17.0%) | 792 (20.2%) | 104 (17.9%) | 122 (25.4%) | 570 (14.4%) | 6342 (15.6%) |
| 65-74 years | 1828 (12.8%) | 2831 (16.3%) | 767 (19.6%) | 132 (22.7%) | 73 (15.2%) | 622 (15.7%) | 6253 (15.4%) |
| 75+ years | 1738 (12.2%) | 1760 (10.2%) | 337 (8.6%) | 35 (6.0%) | 21 (4.4%) | 478 (12.1%) | 4369 (10.8%) |
| *Missing* | 154 | 69 | 10 | 1 | 2 | 51 | 287 |
| **Ethnic group** |  |  |  |  |  |  |  |
| White British | 8637 (60.4%) | 15080 (87.0%) | 3673 (93.7%) | 552 (94.8%) | 445 (92.5%) | 3310 (82.7%) | 31697 (78.0%) |
| White Other | 1166 (8.1%) | 1147 (6.6%) | 133 (3.4%) | 15 (2.6%) | 18 (3.7%) | 327 (8.2%) | 2806 (6.9%) |
| Mixed | 217 (1.5%) | 226 (1.3%) | 47 (1.2%) | 7 (1.2%) | 7 (1.5%) | 53 (1.3%) | 557 (1.4%) |
| Asian | 2901 (20.3%) | 385 (2.2%) | 27 (0.7%) | 4 (0.7%) | 7 (1.5%) | 135 (3.4%) | 3459 (8.5%) |
| Black | 1065 (7.4%) | 372 (2.1%) | 28 (0.7%) | 2 (0.3%) | 2 (0.4%) | 146 (3.6%) | 1615 (4.0%) |
| Other | 321 (2.2%) | 128 (0.7%) | 14 (0.4%) | 2 (0.3%) | 2 (0.4%) | 32 (0.8%) | 499 (1.2%) |
| *Missing* | 92 | 64 | 10 | 0 | 2 | 14 | 182 |
| **Highest qualification** |  |  |  |  |  |  |  |
| No qualification | 3203 (22.4%) | 1692 (9.8%) | 349 (8.9%) | 57 (9.8%) | 69 (14.3%) | 681 (17.1%) | 6051 (14.9%) |
| NVQ <= 3 | 6677 (46.7%) | 8086 (46.7%) | 1796 (45.8%) | 284 (49.0%) | 247 (51.2%) | 2020 (50.7%) | 19110 (47.1%) |
| NVQ4+ (degree) | 3238 (22.7%) | 6405 (37.0%) | 1504 (38.3%) | 199 (34.3%) | 135 (28.0%) | 952 (23.9%) | 12433 (30.6%) |
| Other | 1172 (8.2%) | 1148 (6.6%) | 276 (7.0%) | 40 (6.9%) | 31 (6.4%) | 334 (8.4%) | 3001 (7.4%) |
| *Missing* | 109 | 71 | 7 | 2 | 1 | 30 | 220 |
| **AUDIT-C score** |  |  |  |  |  |  |  |
| 0 | 14399 (100.0%) | 0 (0.0%) | 0 (0.0%) | 0 (0.0%) | 0 (0.0%) | 0 (0.0%) | 14399 (35.4%) |
| 1–4 | 0 (0.0%) | 11895 (68.5%) | 550 (14.0%) | 19 (3.3%) | 19 (4.0%) | 3106 (80.2%) | 15589 (38.4%) |
| 5–7 | 0 (0.0%) | 4418 (25.4%) | 1856 (47.2%) | 136 (23.4%) | 76 (15.8%) | 578 (14.9%) | 7064 (17.4%) |
| 8–12 | 0 (0.0%) | 1064 (6.1%) | 1524 (38.8%) | 425 (73.3%) | 386 (80.2%) | 190 (4.9%) | 3589 (8.8%) |
| *Missing* | 0 | 25 | 2 | 2 | 2 | 143 | 174 |
| **Full AUDIT score** |  |  |  |  |  |  |  |
| 0 | 14050 (97.6%) | 0 (0.0%) | 0 (0.0%) | 0 (0.0%) | 0 (0.0%) | 0 (0.0%) | 14050 (34.6%) |
| 1–7 | 340 (2.4%) | 15221 (87.6%) | 1897 (48.3%) | 104 (18.0%) | 67 (14.0%) | 3489 (90.2%) | 21118 (52.0%) |
| 8–15 | 4 (0.0%) | 2036 (11.7%) | 1846 (47.0%) | 403 (69.6%) | 239 (50.1%) | 322 (8.3%) | 4850 (11.9%) |
| 16–19 | 0 (0.0%) | 93 (0.5%) | 129 (3.3%) | 51 (8.8%) | 73 (15.3%) | 23 (0.6%) | 369 (0.9%) |
| 20–40 | 0 (0.0%) | 23 (0.1%) | 52 (1.3%) | 21 (3.6%) | 98 (20.5%) | 33 (0.9%) | 227 (0.6%) |
| *Missing* | 5 | 29 | 8 | 3 | 6 | 150 | 201 |
| **Favourite drink** |  |  |  |  |  |  |  |
| Beer | 1 (25.0%) | 2218 (39.5%) | 1493 (43.9%) | 259 (46.1%) | 228 (49.2%) | 334 (41.3%) | 4533 (41.7%) |
| Cider | 0 (0.0%) | 318 (5.7%) | 119 (3.5%) | 30 (5.3%) | 42 (9.1%) | 62 (7.7%) | 571 (5.3%) |
| Mixed spirits | 0 (0.0%) | 402 (7.2%) | 165 (4.8%) | 14 (2.5%) | 21 (4.5%) | 71 (8.8%) | 673 (6.2%) |
| Spirits alone | 0 (0.0%) | 473 (8.4%) | 227 (6.7%) | 44 (7.8%) | 49 (10.6%) | 150 (18.5%) | 943 (8.7%) |
| Wine | 1 (25.0%) | 2152 (38.3%) | 1387 (40.8%) | 211 (37.5%) | 122 (26.3%) | 176 (21.8%) | 4049 (37.3%) |
| Other | 2 (50.0%) | 57 (1.0%) | 12 (0.4%) | 4 (0.7%) | 1 (0.2%) | 16 (2.0%) | 92 (0.8%) |
| *Missing* | 14395 | 11782 | 529 | 20 | 20 | 3208 | 29954 |
| **Attempt to cut down** |  |  |  |  |  |  |  |
| No attempt to cut down | 2 (50.0%) | 4302 (76.5%) | 2453 (72.1%) | 400 (71.2%) | 316 (68.1%) | 642 (79.0%) | 8115 (74.7%) |
| Attempt to cut down in last 12 months | 2 (50.0%) | 1319 (23.5%) | 950 (27.9%) | 162 (28.8%) | 148 (31.9%) | 171 (21.0%) | 2752 (25.3%) |
| *Missing* | 14395 | 11781 | 529 | 20 | 19 | 3204 | 29948 |
